# Unveiling Explainable AI in Healthcare: Current Trends, Challenges, and Future Directions

**DOI:** 10.1101/2024.08.10.24311735

**Authors:** Noor A. Aziz, Awais Manzoor, Muhammad Deedahwar Mazhar Qureshi, M. Atif Qureshi, Wael Rashwan

## Abstract

This overview investigates the evolution and current landscape of eXplainable Artificial Intelligence (XAI) in healthcare, highlighting its implications for researchers, technology developers, and policymakers. Following the PRISMA protocol, we analysed 89 publications from January 2000 to June 2024, spanning 19 medical domains, with a focus on Neurology and Cancer as the most studied areas. Various data types are reviewed, including tabular data, medical imaging, and clinical text, offering a comprehensive perspective on XAI applications. Key findings identify significant gaps, such as the limited availability of public datasets, suboptimal data preprocessing techniques, insufficient feature selection and engineering, and the limited utilisation of multiple XAI methods. Additionally, the lack of standardised XAI evaluation metrics and practical obstacles in integrating XAI systems into clinical workflows are emphasised. We provide actionable recommendations, including the design of explainability-centric models, the application of diverse and multiple XAI methods, and the fostering of interdisciplinary collaboration. These strategies aim to guide researchers in building robust AI models, assist technology developers in creating intuitive and user-friendly AI tools, and inform policymakers in establishing effective regulations. Addressing these gaps will promote the development of transparent, reliable, and user-centred AI systems in healthcare, ultimately improving decision-making and patient outcomes.

## 1 INTRODUCTION

Advancements in computer science, particularly in Machine Learning (ML), have played a pivotal role in ushering in the fourth industrial revolution, creating a nexus of interdisciplinary applications that extend beyond traditional boundaries. One of the most significant impact areas is healthcare, where these technologies have gained increasing acceptance and adoption for mission-critical tasks (Jiang et al., 2017).

While integrating AI and ML technologies into healthcare has significantly enhanced decision-making capabilities, it has also highlighted the need for greater transparency. The COVID-19 pandemic, in particular, underscored the importance of these technologies in managing evolving medical knowledge during critical situations (Bullock et al., 2020). AI and ML systems have proven invaluable in disease classification and cancer diagnosis (Esteva et al., 2017). However, their accuracy and precision, though crucial, are not sufficient on their own. It is essential to equip decision-makers with interpretable methods that allow stakeholders to understand, evaluate, and, when necessary, adjust technological decisions (Doshi-Velez & Kim, 2017). The rapid advancements in XAI have made model explainability a central concern, bridging the gap between AI and its application domains, including healthcare, social sciences, and engineering (Holzinger et al., 2017).

This need for explainability is especially critical in clinical decision-making, where healthcare professionals increasingly rely on ML/AI models integrated into standard protocols (Loh et al., 2022). Implementing explainable AI within healthcare systems is crucial to ensuring informed, ethical, and effective patient care as healthcare embraces digital transformation (Band et al., 2023).

XAI has thus emerged as a vital area of research, focusing on making AI models more interpretable and transparent without compromising performance. The significance of XAI goes beyond theory; it directly impacts the usability and reliability of AI systems in real-world healthcare applications (Allgaier et al., 2023). Consequently, XAI has attracted significant attention from diverse stakeholders, including healthcare AI researchers, technology developers, and policymakers. These stakeholders play critical roles in advancing the field, from developing new XAI methods and translating them into practical applications to ensuring that these technologies align with ethical standards and regulatory requirements (Topol, 2019). While these stakeholders drive the development and implementation of XAI, the primary beneficiaries are healthcare professionals, who rely on these systems to make informed decisions that profoundly impact patient outcomes.

### 1.1 Need for Explainable AI in Healthcare

Decision support tools are indispensable in modern healthcare, aiding doctors, nurses, and pharmacists in making informed patient care decisions (Wasylewicz & Scheepers-Hoeks, 2019). These systems range from basic information providers, such as pharmacy databases (Alanazi et al., 2018), to advanced algorithms that recommend personalised treatments (Kronenfeld et al., 2013). Some tools operate automatically, offering immediate guidance (Papadopoulos et al., 2022), while others depend on manual input, such as clinical guidelines (Feder et al., 1999).

The digital transformation of healthcare, particularly with the advent of Electronic Health Records (EHRs), has led to a significant increase in data generation. EHRs capture extensive patient health information as a digital alternative to traditional paper records (Hersh, 2004). This wealth of data fuels the development of advanced decision support systems that empower providers to make informed decisions based on specific patient conditions (DesRoches et al., 2008).

These systems have demonstrated the potential to reduce errors and improve patient outcomes, although their presence alone does not guarantee better care. Research indicates a notable decrease in decision-making errors (Magrabi et al., 2016), but the effectiveness of these systems largely depends on their quality and the accuracy of the data they utilise.

The value of decision support systems is particularly evident in situations that demand quick, informed decisions. By providing clinicians with immediate access to relevant patient data and the latest medical insights, these tools enable healthcare professionals to manage complex information, ensuring timely and well-informed decisions (Vaishya et al., 2020). The primary aim of AI in healthcare is to enhance patient care through evidence-based clinical decision-making.

XAI is crucial in this context to ensure the transparency and interpretability of these tools (Guidotti et al., 2018). Integrating XAI into healthcare enhances the accuracy of AI decisions and makes them understandable to clinicians. This transparency is vital for building trust with healthcare providers, enabling effective scrutiny and validation of system recommendations (Tjoa & Guan, 2020).

### 1.2 Motivation for the work

The integration of AI in healthcare has introduced advanced decision-making tools with the potential to transform patient care. However, the widespread adoption of these technologies is hindered by their lack of transparency and interpretability, often referred to as the “black-box” problem. This issue is particularly critical in healthcare, where clinical decisions must be justifiable and understandable to healthcare providers, patients, and regulatory bodies.

XAI methods have emerged as a potential solution to this challenge, aiming to make AI models more interpretable while maintaining high performance. Despite the growing body of research on XAI, the literature remains fragmented and often lacks a comprehensive approach. Many studies focus on specific use cases or methods without addressing XAI’s broader applicability and effectiveness across different healthcare domains.

For instance, a survey by Antoniadi et al. (2021) touches on XAI methods in healthcare but does not focus on specific ML models or application areas. Similarly, Xu et al. (2023) discusses AI in healthcare from a technological perspective but lacks detailed insights into machine learning models, XAI methods, and datasets. Ali et al. (2023) focus on XAI methods and datasets but offer limited discussion on machine learning models and healthcare applications. Moreover, the review by Prentzas et al. (2023) explores the implementation of XAI from a physician’s perspective, yet it lacks coverage of ML models and dataset characteristics.

Additionally, a review by Loh et al. (2022) provides a high-level overview of XAI methods but does not delve deeply into specific ML models or application areas, leaving a gap in the practical understanding of these methods. Band et al. (2023) focus on image-based datasets for XAI but neglect to discuss free-text and tabular data, which are also crucial in healthcare applications. Furthermore, Allgaier et al. (2023) explores the taxonomy of XAI methods but provides limited discussion on application areas, datasets, and ML models, which are essential for a comprehensive understanding of XAI in healthcare.

As summarised in Table 2, existing studies show substantial variability in methodological rigour regarding XAI methods and ML models. Notably, none have reviewed multiple XAI methods in a single study, and many lack comprehensive datasets or detailed discussions on various application areas.

**TABLE 1.** List of abbreviations and their descriptions.

| Abbreviation | Description | Abbreviation | Description |
| --- | --- | --- | --- |
| RNN | Recurrent Neural Network | GRU | Gated Recurrent Unit |
| LSTM | Long Short-Term Memory | CNN | Convolutional Neural Network |
| LR | Logistic Regression | SVM | Support Vector Machine |
| RF | Random Forest | XGB | Extreme Gradient Boost |
| MLP | Multi-Layer Perceptron | PFI | Permutation Feature Importance |
| DT | Decision Trees | KNN | k-Nearest Neighbors |
| DNN | Deep Neural Network | ELI5 | Explain Like I am 5 |
| ICE | Individual Conditional Expectation | GBDT | Gradient Boosted Decision Trees |
| SVC | Support Vector Classification | GB | Gradient Boosting |
| LGB | Light Gradient Boosting Machine | CDSS | Clinical Decision Support System |
| EHR | Electronic Health Records | LGA | Large for Gestational Age |
| LIME | Local Interpretable Model-agnostic Explanations | AGRAD | Attention Gradient |
| LORE | LOcal Rule-based Explanations | LRP | Layer-wise Relevance Propagation |
| GradCAM | Gradient-weighted Class Activation Mapping | PDP | Partial dependence plots |
| SHAP | Shapley Additive Explanations | CIU | Contextual Importance and Utility |
| FI | Feature Importance | BoCSor | Boundary Crossing Solo Ratio |
| ABELE | Anchors Basic Explanation Linked-Examples Enhanced Explanation |  |  |

**TABLE 2.** Summary of Related Literature Reviews.

| Ref. | Published | Duration | Application Area | Dataset Characteristics | ML Models | XAI Methods | Multiple XAI Methods | Remarks |
| --- | --- | --- | --- | --- | --- | --- | --- | --- |
| Antoniadi et al. (2021) | 2021 | 2008-2020 | - | - | - | Low | - | Touches upon XAI methods in CDSS without focusing on specific ML models. |
| Loh et al. (2022) | 2022 | 2011-2022 | Low | - | Low | High | - | Focuses on XAI methods with limited discussion on ML models and application areas. |
| Xu et al. (2023) | 2023 | 2011-2020 | - | - | - | Low | - | Discusses CDSS from a technological and medical perspective, with no detailed discussion of ML models, XAI methods, and datasets. |
| Ali et al. (2023) | 2023 | 2018-2022 | Low | Medium | Low | Medium | - | Focuses on XAI methods and the datasets for CDSS, with a limited discussion on ML models and application areas and no discussion of using multiple XAI methods in a single study. |
| Prentzas et al. (2023) | 2023 | upto Oct 2022 | - | - | - | High | - | Reviews the implementation of XAI methods and their challenges from a physician's perspective, with no discussion of ML models, datasets, and applications. |
| Band et al. (2023) | 2023 | 2019-2022 | Medium | Medium | Medium | High | - | Focuses on different XAI methods mostly on image-bases datasets, and lacks a discussion on free-text and tabular datasets. |
| Allgaier et al. (2023) | 2023 | 2020-2022 | - | Low | Low | Medium | - | Focuses on the taxonomy of XAI methods in the medical field, lacking discussion on application areas, with limited discussion on datasets and ML models. |
| S. Y. Kim et al. (2024) | 2024 | 2000-2024 | Low | Low | Low | Medium | - | Covers all areas without delving deeply into any single aspect. |
| Evans et al. (2024) | 2024 | 1998-2024 | - | - | - | - | Low | Only discusses from the perspective of user trust and acceptability. |
| *This review* | 2024 | 2003-2023 | High | High | High | High | High | Provides a comprehensive analysis of XAI methods applied to different ML model categories across various healthcare application areas, detailing dataset characteristics and lists tradeoffs between XAI methods used. |

This review is motivated by the need to address these gaps by providing a comparative analysis of XAI methods in healthcare. Specifically, it aims to assess the effectiveness, applicability, and practical constraints of different XAI approaches across various clinical settings. By offering a broader evaluation and insights into real-world applications, this review seeks to provide actionable guidance to support developing and adopting transparent, reliable, and user-friendly AI systems in healthcare.

### 1.3 Research Questions

This review is guided by the following key research questions to address the existing gaps in XAI research within healthcare:

1. **Which healthcare domains have adopted XAI methods, and what types of datasets are commonly used?** This question explores the most widely implemented XAI methods across clinical areas and assesses the diversity of datasets employed in these studies.
2. **What are the prevailing trends and most effective machine learning models in explainable healthcare systems?** This explores which ML models are most frequently paired with XAI methods and how effectively they enhance interpretability in healthcare contexts.
3. **What are the most commonly used XAI methods in healthcare, and how do they enhance the interpretability of machine learning models?** This focuses on evaluating specific XAI techniques and their role in improving decision-making.
4. **How do multiple XAI methods contribute to explainability in healthcare decision-making processes?** Given the complexity of healthcare decisions, this examines the benefits of employing multiple XAI methods to provide more robust interpretability.

### 1.4 Contributions of This Research

The primary contributions of this research are as follows (Table 2 provides a comparison with related literature reviews):

1. **Comprehensive Analysis of XAI Adoption:** This review offers an in-depth analysis of XAI applications across healthcare, identifying both well-explored and underrepresented areas.
2. **Evaluation of Datasets, ML Models and XAI Methods:** The review assesses the characteristics of datasets and machine learning models used in XAI studies, offering insights into their generalisability.
3. **Insights into Multiple XAI Methods:** We evaluate the use of multiple XAI techniques, emphasising the importance of combining methods to improve model interpretability.
4. **Identification of Research Gaps:** This review highlights critical gaps in the current XAI landscape and provides actionable recommendations for future research.

### 1.5 Organisation of the Article

The remainder of this article is structured as follows: Section 2 outlines the methodology, including the PRISMA protocol and study selection criteria. Section 3 reviews XAI applications across various healthcare domains, focusing on different application areas and the datasets used. Section 4 discusses the predictive models commonly employed in healthcare and their integration with XAI methods. Section 5 provides an analysis of various XAI techniques and their role in enhancing model interpretability and decision-making processes. Section 6 examines studies utilising multiple XAI methods to combine the strengths of different approaches for better interpretability. Section 7 highlights open research gaps in the field and offers actionable recommendations for future work. Finally, Section 8 summarises the key findings of this review and presents concluding thoughts on the future direction of XAI in healthcare.

To provide a clear and concise visual overview of the article’s structure and its relationships, we have included a Figure 1. This figure illustrates the logical connections between different sections and their contributions to the overall narrative of the article.

**FIGURE 1.**
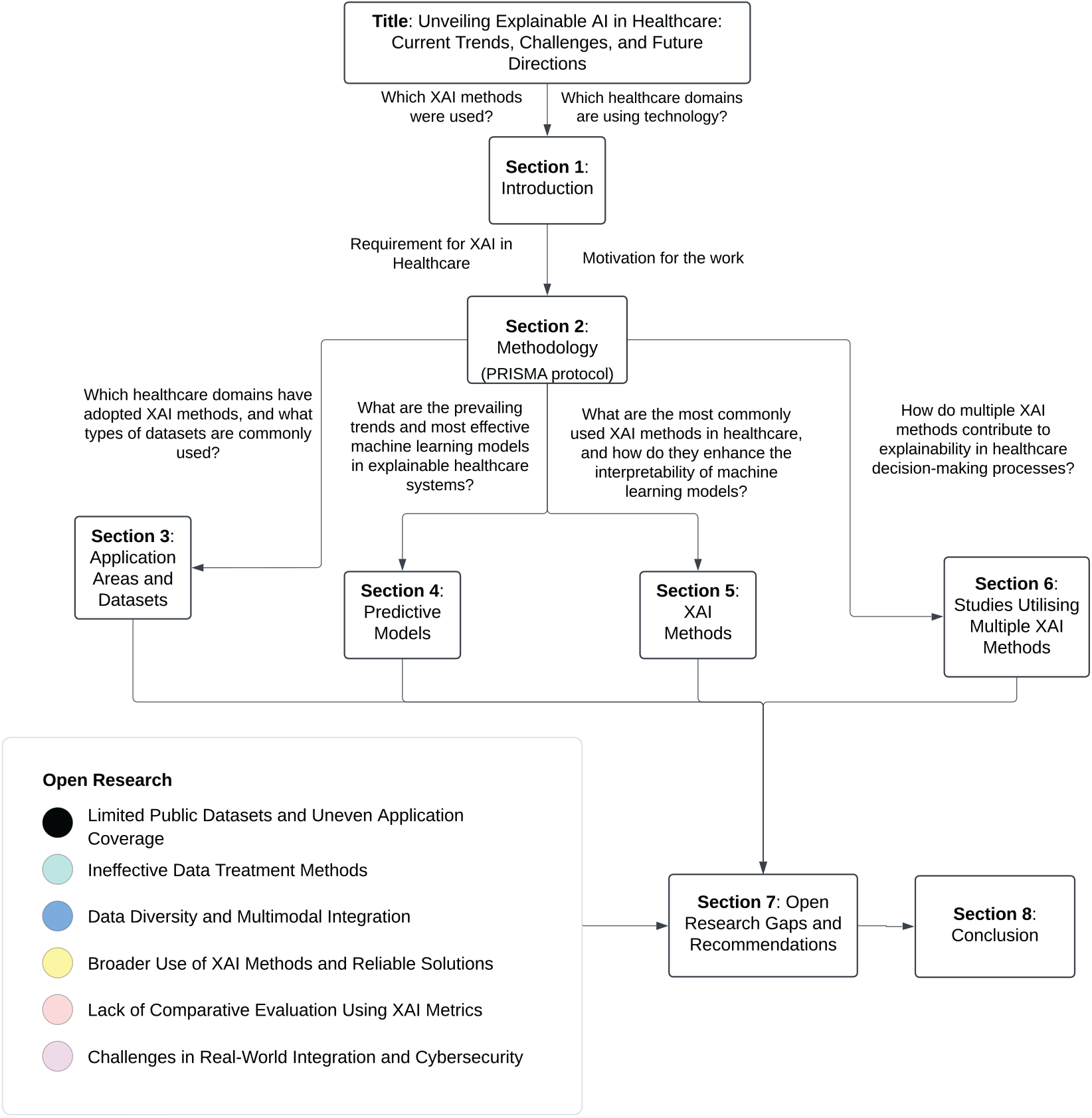
The figure outlines the article’s structure, beginning with the Introduction and moving through the Methodology, Applications, Predictive Models, XAI Methods, Studies Utilising Multiple XAI Methods, and concluding with Research Gaps and Recommendations.

## 2 METHODOLOGY

To conduct this review, we followed a rigorous methodology, drawing upon the established literature review framework (Keele et al., 2007) and aligning our protocols with PRISMA-P guidelines (Moher et al., 2015) to ensure a comprehensive approach. Figure 2 illustrates the detailed implementation of the PRISMA-P protocol tailored to our review objectives.

**FIGURE 2.**
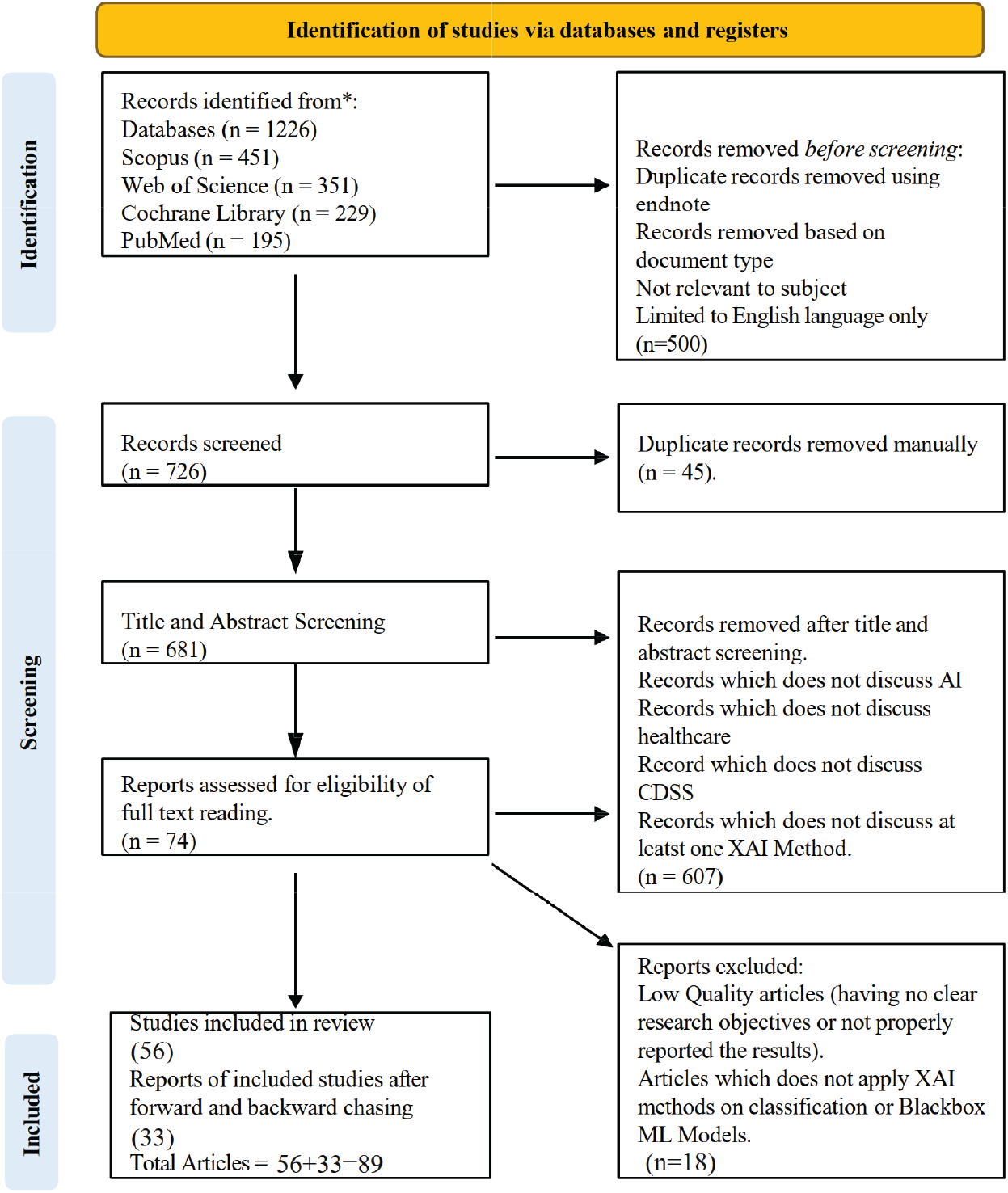
PRISMA Diagram

The following sections outline the review methodology, including the eligibility criteria, identification of relevant information sources, formulation of the search strategy, and the screening process undertaken to curate the final set of articles for analysis.

### 2.1 Eligibility Criteria

The eligibility criteria for this review were carefully defined to ensure the inclusion of studies that offer meaningful insights into applying XAI in healthcare. Studies were considered eligible if they met the following conditions:

1. **Time Frame:** The study was published between January 2000 and June 2024, covering the period of integrating AI and XAI methods into healthcare technologies.
2. **Healthcare Focus:** The study specifically addressed the application of XAI techniques within healthcare settings, including but not limited to clinical decision-making, diagnostics, treatment planning, and patient management.
3. **AI and XAI Methods:** The study utilised or evaluated AI models where XAI methods were applied to enhance interpretability and transparency. This includes studies discussing the development, implementation, or assessment of XAI techniques in various healthcare contexts.
4. **Data Types:** The study used relevant healthcare data types, such as tabular data, medical imaging, clinical text, or sensor data, ensuring the analysis was grounded in practical, real-world applications.
5. **Peer-Reviewed Articles:** Only peer-reviewed articles were included to ensure the credibility and scientific rigour of the studies.
6. **Language:** The study was available in English, ensuring accessibility and comprehensibility for researchers and practitioners.

Studies that did not meet these criteria, such as those focused on non-healthcare applications, lacking XAI components, or not peer-reviewed, were excluded from the review.

### 2.2 Databases

We utilised multiple databases to gather information, including Scopus, Web of Science, Cochrane Library, IEEE Explore, PubMed, and Science Direct. Although we received almost identical outcomes from different databases, we focused on four primary databases to eliminate redundancy and duplicate publications: Scopus, Web of Science, PubMed, and Cochrane Library.

Scopus and Web of Science are extensive databases encompassing various research topics and publications, making them reliable literature sources across various fields. Meanwhile, PubMed and Cochrane Library are essential sources of knowledge for medical and biomedical studies. PubMed covers advancements in medical expertise, while Cochrane Library is a valuable resource for clinical studies examining clinical practices, implementations, and outcomes. Although we tested our search terms on IEEE Explore, it yielded insufficient results. We also found that Science Direct produced nearly identical outcomes to Scopus and Web of Science. However, we focused on these two databases due to the broad range of multidisciplinary coverage offered by Scopus and Web of Science.

### 2.3 Search Strategy

The search strategy was designed to capture a broad range of studies related to XAI in healthcare, covering the period from January 2000 to June 2024. We focused on two major concepts that define our research topic: “clinical decision support systems” and “explainable artificial intelligence.” The search strategy was divided into three parts based on these concepts. The first part focused on explainability, covering all relevant synonyms and related terms using the OR operator. The second part addressed artificial intelligence, encompassing publications related to AI, while the third part represented clinical decision support systems, incorporating synonyms and related terms. These three parts were combined using the AND operator (see Table 3).

**TABLE 3.** Search Terms.

| Search Terms | Operator | Search Terms | Operator | Search Terms |
| --- | --- | --- | --- | --- |
| ( explainable OR interpretable OR transparent OR accountable OR human-interpretable OR human-centered ) | AND | ( AI OR ML OR Deep Learning OR machine AND learning OR deep AND learning OR artificial AND intelligence OR computational AND intelligence ) OR XAI ) | AND | (( ( health* OR *medical OR clinical ) AND decision* AND ( support OR making OR aids ) AND system* ) OR cdss OR cds ) ) |

After designing the search terms, we applied them across the selected databases to the topic title, abstract, and keyword fields. The search query was adapted according to each database’s search policy (see Table 1 in Appendix). The search was conducted within the timeframe of 01-Jan-2000 to 31-Dec-2023, yielding a total of 1226 research articles^‡^. Articles from 2024 published until June were also included, adding 19 articles^*§*^ to the total.

### 2.4 Screening of Studies

We undertook several steps to ensure the inclusion of high-quality, relevant articles based on our inclusion and exclusion criteria (see Table 4). Initially, we applied filters based on document type, subject matter, and language (English only) to limit our search results to peer-reviewed journal articles focused on XAI in healthcare. After applying these filters, we used Endnote reference management software to detect and remove duplicate articles and manually removed any remaining duplicates. This process left us with 681 articles (see Figure 2). Further screening was conducted in two stages: title and abstract screening, followed by a full-text review. To ensure the rigour and objectivity of the screening process, multiple reviewers independently assessed the relevance of each study.

**TABLE 4.** Inclusion and Exclusion Criteria.

| Inclusion | Exclusion |
| --- | --- |
| Discusses XAI in healthcare | Lacks focus on healthcare and CDSS |
| Discusses XAI and CDSS | Not written in English |
| Utilises ML models and XAI methods with clinical datasets | Duplicate articles |
|  | Involves fuzzy classifiers |
|  | Philosophical, Review, Conference, or Discussion studies |

#### Title and Abstract Screening

In the first stage, two author reviewers screened the titles and abstracts of all identified records. Each reviewer consistently applied the inclusion and exclusion criteria to determine whether the study warranted further examination. If there were any discrepancies between the reviewers regarding the eligibility of a study, these were resolved through discussion and consensus. In cases where consensus could not be reached, a third reviewer^¶^ was consulted to make the final decision. After this stage, 74 articles met our criteria and were subjected to a full-text review.

#### Full-Text Review

In the second stage, the full texts of the studies that passed the initial screening were retrieved and reviewed in detail by the same two reviewers. The inclusion and exclusion criteria were applied more rigorously to ensure that only studies meeting all the requirements were included in the final analysis. We only included articles with clear research objectives, correctly reported results, discussed the application of the XAI method on classification or black-boxed machine learning models, and at least one explanation method applied to clinical data. Any disagreements during the full-text review were similarly resolved through discussion or consulting a third reviewer. Ultimately, we were left with 56 articles that matched our inclusion criteria.

#### Reference Chasing

To ensure no relevant articles were missed, we also conducted forward and backward reference chasing. This resulted in the inclusion of 33 additional articles that met our inclusion criteria, 19 of which were published from January to June 2024.

#### Consistency and Reliability

To enhance the reliability of the screening process, the reviewers underwent a calibration exercise before beginning the screening. This involved reviewing a sample set of studies together and discussing the application of the criteria to ensure consistency in their approach. This activity was overseen by the senior authors^#^.

By utilising multiple reviewers and adhering to these best practices, we minimised the risk of bias in the study selection process, ensuring a comprehensive and objective review of the literature on XAI in healthcare.

### 2.5 Data Extraction and Synthesis

This review includes articles published from January 2000 to June 2024. After applying the search criteria and filtering studies based on inclusion and exclusion criteria, we found that most studies were published in recent years. Specifically, seven articles were published in 2020, nine in 2021, fourteen in 2022, forty in 2023, and nineteen articles published by June 2024. The gradual increase in studies over recent years indicates a growing interest from researchers and stakeholders in applying artificial intelligence within the healthcare domain. Consequently, it is an opportune time to evaluate the current state of XAI in CDSS, understand the existing challenges, and provide insights for future implementation and research.

The studies selected for this review underwent a detailed data extraction process to gather important information. A standardised data extraction form was used to collect data on study characteristics, including study design, sample size, XAI methods used, ML models implemented, datasets’ characteristics, and the specific healthcare applications addressed. This thorough extraction aimed to ensure a comprehensive understanding of how XAI is used within CDSS and identify any prevalent trends or research gaps.

The extracted data were then synthesised to provide a comprehensive overview of the current state of XAI in healthcare. The synthesis focused on identifying trends, evaluating the effectiveness of various XAI methods, and highlighting gaps in the literature. Comparative analysis was also performed to assess the strengths and limitations of different XAI approaches.

### 2.6 Limitations of the Methodology

While this review followed a rigorous methodology, certain limitations should be acknowledged. The reliance on English-language articles may have excluded relevant studies in other languages. Additionally, the focus on peer-reviewed literature may have excluded emerging research that has not yet undergone formal peer review. Despite these limitations, the methodology employed in this review provides a comprehensive and reliable analysis of the current landscape of XAI in healthcare.

## 3 APPLICATION AREAS AND DATASETS

This section presents a descriptive analysis of the application areas where XAI methods have been utilised in healthcare, along with the types and characteristics of datasets employed in these studies. The descriptive statistics cover 89 studies across 19 distinct medical domains, such as Neurology, Cancer, Cardiovascular Diseases, COVID-19, Diabetes, and others, which are summarised in Table 5.

**TABLE 5.**
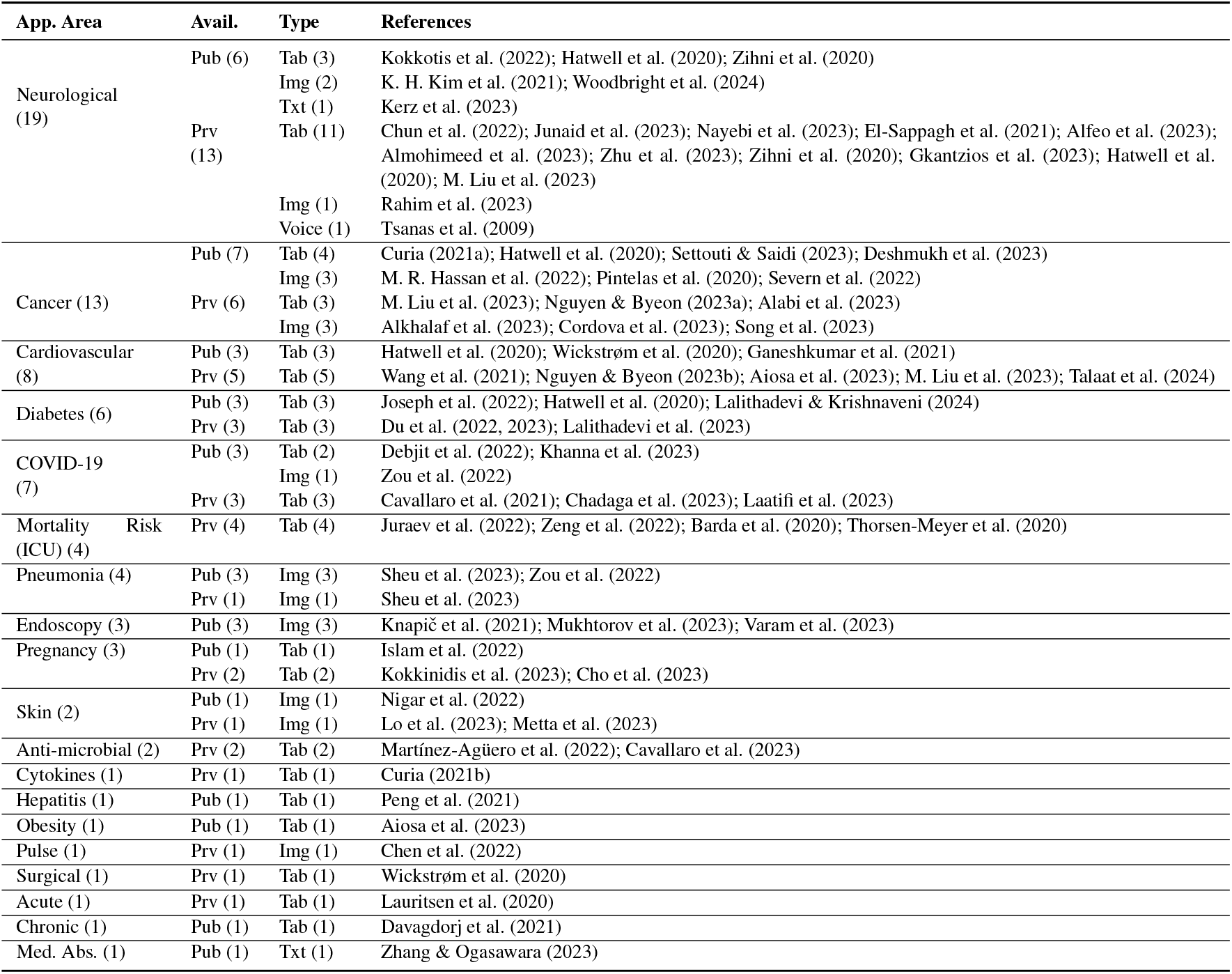
Overview of datasets and application areas.

Figure 3 provides a comprehensive visualisation of the proportional research focus across various healthcare domains. The chart highlights Neurology and Cancer as the most extensively studied areas, followed by Cardiovascular Diseases and Diabetes. This visualisation complements the discussion in this section, offering an intuitive understanding of the distribution of XAI applications within healthcare. Also, Figure 4 provides a visual representation of the dataset distribution across different application areas, highlighting the diversity and characteristics of datasets used in XAI applications.

**FIGURE 3.**
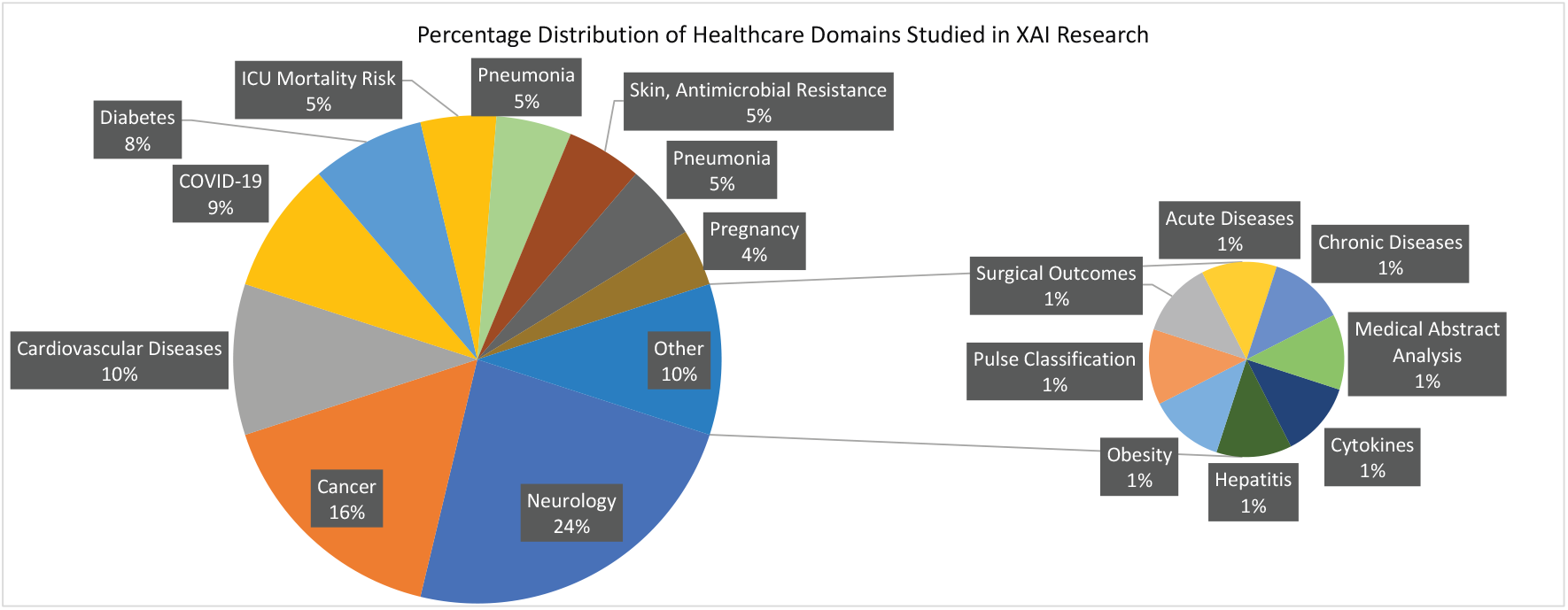
Healthcare Domains in XAI Research: Percentage Distribution

**FIGURE 4.**
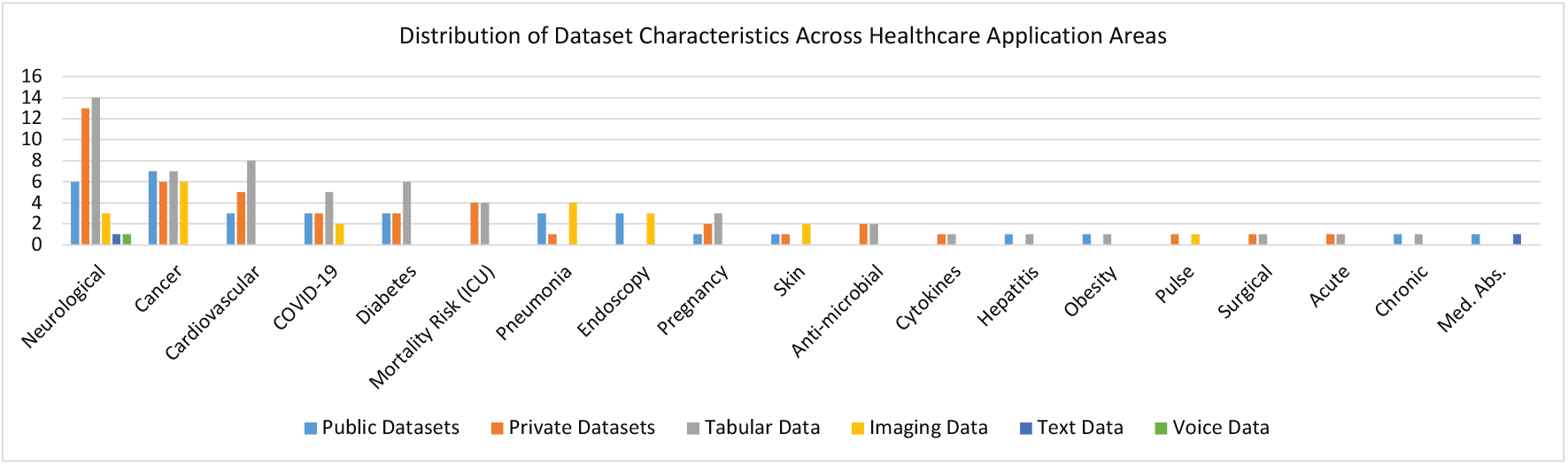
Distribution of Dataset Characteristics in Healthcare Applications.

The discussion begins with the most prominent categories, followed by a combined subsection addressing less frequently studied areas. Lastly, the dataset challenges subsection outlines key issues such as data imbalance, missing values, and reliance on private datasets, which affect the generalisability and robustness of XAI models in healthcare. Detailed descriptions of relevant datasets, including their availability and types (e.g., tabular, image, text), can be found in the Appendix (Tables 2 and 3); readers may skip these tables if desired.

### 3.1 Neurological

Neurological conditions represent the most extensively researched category, with 19 studies dedicated to diagnosis and assessment (see Table 5). These studies encompass a variety of neurological disorders, including Alzheimer’s disease (El-Sappagh et al., 2021; Almohimeed et al., 2023; Rahim et al., 2023; Woodbright et al., 2024), stroke (Kokkotis et al., 2022; Zihni et al., 2020; Gkantzios et al., 2023; Mridha et al., 2023), dementia (Chun et al., 2022), Parkinson’s disease (Junaid et al., 2023; Ducange et al., 2024), cerebrovascular issues (K. H. Kim et al., 2021), brain injuries (Nayebi et al., 2023), depressive disorder (Zhu et al., 2023), language behaviour-based mental health issues (Kerz et al., 2023), and brain connectivity networks (Alfeo et al., 2023). The studies utilised 13 private and six public datasets, incorporating various data types, including tabular (14), image (3), text data (1), and voice (1). The limited use of public datasets (6 studies) and image data (3 studies) suggests challenges in data accessibility and a potential underutilisation of imaging despite its importance in neurological diagnosis. This highlights the need for more publicly available and diverse datasets to improve the robustness and generalisability of AI models in this field. These datasets often exhibit imbalances and contain missing values, necessitating rigorous preprocessing to ensure robust analysis and accurate model performance.

### 3.2 Cancer

Cancer detection and diagnosis represent a significant focus area, with 13 studies covering various types of cancer, including cervical (Curia, 2021a), liver (M. Liu et al., 2023), multiple myeloma (Settouti & Saidi, 2023), prostate (M. R. Hassan et al., 2022), lung (Nguyen & Byeon, 2023a; M. Liu et al., 2023; Wani et al., 2024), glioma brain (Pintelas et al., 2020; Severn et al., 2022), breast (Cordova et al., 2023; Hatwell et al., 2020; M. Liu et al., 2023), nasopharyngeal (Alabi et al., 2023), colorectal (Alkhalaf et al., 2023), and tumour (Song et al., 2023). The cancer research studies show a balanced use of public (7) and private (6) datasets, aiding data accessibility and validation. The studies predominantly use tabular (7) and image data (6), highlighting the importance of structured information and medical imaging in cancer diagnosis and treatment. This balance underscores the critical role of explainable AI in interpreting complex data, which is crucial for advancing cancer research. Like neurological datasets, cancer datasets often exhibit imbalances and missing values, necessitating appropriate preprocessing measures.

### 3.3 Cardiovascular

Seven studies centred on cardiovascular conditions, including cardiac arrest (Nguyen & Byeon, 2023b), heart failure with coronary heart disease (Wang et al., 2021), comorbidity (Aiosa et al., 2023), hypertensive heart disease (M. Liu et al., 2023), myocardial infarction (Wickstrøm et al., 2020), assessment via electrocardiograms (Ganeshkumar et al., 2021; Talaat et al., 2024), and cardiography (Hatwell et al., 2020). All Cardiovascular research studies use tabular datasets (8), mainly private data (5). This reliance highlights challenges in data accessibility and a gap in utilising diverse data types, such as text and imaging. Additionally, imbalances and missing values in datasets point to the need for improved preprocessing. Expanding data diversity and accessibility could enhance the effectiveness of the AI model in this field.

### 3.4 COVID-19 and Diabetes

Seven studies each focused on COVID-19 and diabetes. The COVID-19 studies investigated areas such as COVID-19 prediction (Debjit et al., 2022; M. M. Hassan et al., 2024), ICU admissions (Cavallaro et al., 2021), diagnosis of influenza-like illness (Chadaga et al., 2023), triage prediction systems (Khanna et al., 2023), severity risk assessment (Laatifi et al., 2023), severe community-acquired pneumonia, and respiratory infections (Zou et al., 2022). The diabetes studies examined the prediction of large gestational age (LGA) in overweight and obese female patients (Du et al., 2022, 2023; Lalithadevi & Krishnaveni, 2024), general classification and prediction of diabetes (Joseph et al., 2022), and diabetic retinopathy in type 2 diabetes patients (Lalithadevi et al., 2023; Hatwell et al., 2020). The datasets for both categories are primarily tabular, except for two image datasets in the COVID-19 category. Each category includes three public and three private datasets, with issues related to missing values and outliers. The diabetes datasets showed less imbalance than the COVID-19 datasets, with one notable exception being a highly imbalanced dataset.

### 3.5 Mortality Risk Prediction (ICU)

Four studies focused on mortality risk prediction in ICU settings, including a qualitative analysis in pediatric intensive care units (Barda et al., 2020), overall mortality risk prediction (Thorsen-Meyer et al., 2020; Juraev et al., 2022), and assessing the risk of extubation failure in ICU patients undergoing ventilation (Zeng et al., 2022). All datasets used in these studies are private, comprising patient vitals, hospital records, and laboratory tests. These datasets are frequently imbalanced and contain missing values and outliers, necessitating thorough preprocessing to ensure accurate model predictions.

### 3.6 Pneumonia, Pregnancy, and Endoscopy

Three studies each focused on pregnancy (Islam et al., 2022; Kokkinidis et al., 2023; Cho et al., 2023) and endoscopy (Knapič et al., 2021; Mukhtorov et al., 2023; Varam et al., 2023), while two studies addressed pneumonia (Sheu et al., 2023; Zou et al., 2022). In pregnancy, studies predicted preterm births (Kokkinidis et al., 2023), extrauterine growth restriction (Cho et al., 2023), and anticipated cesarean delivery outcomes (Islam et al., 2022). Endoscopy and pneumonia studies primarily used publicly available image-based datasets, except for one private pneumonia dataset. Two of the endoscopy datasets are multiclass, and one is highly imbalanced. The pregnancy category datasets are tabular, imbalanced, and contain missing values.

### 3.7 Rare Applications

Less frequently studied application areas include skin lesion classification (Nigar et al., 2022), skin vascular wound images (Lo et al., 2023), antimicrobial resistance (Cavallaro et al., 2023; Peng et al., 2021), hepatitis liver disease (Peng et al., 2021), pulse wave classification (Chen et al., 2022), obesity (Aiosa et al., 2023), surgical outcomes (Wickstrøm et al., 2020), chronic disease (Davagdorj et al., 2021), and acute disease (Lauritsen et al., 2020). In the antimicrobial domain, early detection of drug resistance was investigated by Martínez-Agüero et al. (2022), while antimicrobial stewardship was examined by Cavallaro et al. (2023). These studies employed tabular and image datasets, sourced from a mix of public and private origins, often facing challenges of imbalance and missing data. Despite their relative rarity, these applications demonstrate the versatility of XAI methods in addressing a wide range of medical conditions.

### 3.8 Dataset Challenges

As discussed above, tabular datasets are often imbalanced, with missing values and noise, necessitating careful preprocessing and balancing to improve results. Common approaches to handling missing values (when more than 30% is missing) include dropping data (Junaid et al., 2023; Almohimeed et al., 2023; Kokkotis et al., 2022; Zhu et al., 2023; Zihni et al., 2020; Hatwell et al., 2020; M. Liu et al., 2023; Nguyen & Byeon, 2023a; Aiosa et al., 2023; Du et al., 2022, 2023; Kokkinidis et al., 2023; Davagdorj et al., 2021; Curia, 2021b), mean, median, or mode imputation (Mridha et al., 2023; Du et al., 2022, 2023; Chadaga et al., 2023; Zeng et al., 2022; Cho et al., 2023; Curia, 2021b; Junaid et al., 2023; Almohimeed et al., 2023; Joseph et al., 2022; Lalithadevi & Krishnaveni, 2024), KNN interpolation (El-Sappagh et al., 2021; M. Liu et al., 2023; Islam et al., 2022), and last observation carried forward (Thorsen-Meyer et al., 2020; Zeng et al., 2022). However, as (Lalithadevi & Krishnaveni, 2024) noted, the mean is sensitive to noise and can result in incorrect imputations. Other methods include iterative imputation (Debjit et al., 2022; Laatifi et al., 2023), baseline wander removal, and power line interference removal (Ganeshkumar et al., 2021), and forward imputation (Nayebi et al., 2023).

For data balancing, SMOTE (Almohimeed et al., 2023; Settouti & Saidi, 2023; Chadaga et al., 2023; Kokkinidis et al., 2023; Juraev et al., 2022) and its variations, such as SMOTE-ENN (Wang et al., 2021; Nguyen & Byeon, 2023b), borderline SMOTE (Khanna et al., 2023), SMOTE-Tomek (Khanna et al., 2023), and SMOTE-NC (Juraev et al., 2022), are the most common methods applied. Other techniques include random subsampling (Zihni et al., 2020; Kokkinidis et al., 2023), under-sampling (Kokkinidis et al., 2023), and ADASYN (Khanna et al., 2023; Islam et al., 2022).

Across these application areas, tabular datasets frequently suffer from imbalances and missing values, complicating data analysis and model training. The reliance on private datasets, particularly in neurological, ICU, and antimicrobial settings, poses significant challenges to replicability and comparative analysis. For instance, 13 out of 17 datasets are private in the neurological category. Similarly, all datasets related to mortality risk prediction, antimicrobial resistance, and pulse wave classification are private. This high percentage of private datasetsover 50% in most categoriesrepresents a substantial barrier to the broader application and validation of XAI models across diverse healthcare settings.

Moreover, the variability in data types and the specific challenges associated with each (e.g., image versus tabular data) suggest that XAI methods must be tailored to the characteristics of the dataset in use. For instance, image datasets may require methods focusing on spatial features, while tabular data might benefit more from techniques emphasising feature importance and interpretability. Therefore, further research is needed to explore how XAI techniques can be optimised for different health-care data types and effectively integrated into clinical workflows. Readers are encouraged to refer to Tables 2 and 3 in the Appendix for a detailed view of the datasets.

## 4 PREDICTIVE MODELS

This section provides an overview of the predictive models employed in the reviewed studies, categorising them by application area and model type. The choice of machine learning (ML) models is critical, as it directly impacts the performance, interpretability, and explainability of AI systems in clinical settings. ML models are broadly classified into interpretable and non-interpretable categories. Interpretable models, such as Logistic Regression (LR), Decision Trees, and Naive Bayes, are simple and transparent, making their predictions easily understandable. In contrast, non-interpretable models, including non-linear SVMs, ensembles, and deep learning models, are more complex and require explainer methods to clarify or explain their decision-making processes.

Figure 6 illustrates the frequency of predictive models used across different healthcare applications, highlighting the dominance of CNNs and Random Forest models in the studies reviewed. This visualisation provides a clear understanding of the popularity and applicability of these models.

This section begins with an overview of the models used across various healthcare domains. It then discusses model selection based on application area, examining how different clinical contexts influence the choice of predictive models. The section further explores recent trends and insights from 2020 to 2023 concerning model adoption. Finally, it addresses the challenges associated with implementing these models, particularly regarding interpretability and integration into clinical workflows.

### 4.1 Overview of Models Used

The reviewed studies employed various predictive models, with a notable preference for ensembles of tree-based models (e.g., Random Forest, Gradient Boosting Machines), SVMs, neural networks, and logistic regression. These models were often selected for their ability to handle specific characteristics of healthcare data, such as high dimensionality, non-linearity, and class imbalances.

### 4.2 Model Selection Based on Application Area

The selection of predictive models varied according to the application area:

- **Neurological Disorders**: Tree-based models, such as Random Forest and Gradient Boosting Machines, were predominantly used due to their ability to handle complex interactions between features in tabular datasets. These models also facilitated the application of feature importance-based XAI methods, which are crucial for understanding the factors contributing to neurological conditions.
- **Cancer Diagnosis**: Neural networks, particularly CNNs, were extensively employed in image-based cancer detection tasks, while logistic regression and SVMs were applied to tabular data scenarios. The use of CNNs in imaging studies underscores the importance of spatial feature extraction in cancer diagnosis, although these models required post-hoc XAI methods to enhance transparency.
- **Cardiovascular Diseases**: Studies in this area often employed a mix of tree-based models and neural networks. SVMs were also common, particularly in studies focused on electrocardiogram (ECG) analysis, where the separation of signal classes was critical. These models were selected for their ability to manage cardiovascular data’s high-dimensional, time-series nature. Additionally, Ghadekar et al. (2024) investigate ensemble tree-based and interpretable models to examine the link between cardiovascular disease and diabetes, while Talaat et al. (2024) employs complex attention-based neural networks.
- **COVID-19 and Diabetes**: For COVID-19 prediction and diabetes classification, a combination of tree-based models and logistic regression was typically employed. These models were chosen for their balance between performance and interpretability, with XAI methods like SHAP frequently applied to provide insights into model decisions.
- **Mortality Risk Prediction (ICU)**: In ICU settings, where timely and accurate predictions are critical, studies favoured tree-based models for their interpretability and ability to handle heterogeneous data types, including patient vitals and clinical records. Logistic regression was also used for its simplicity and ease of integration into clinical workflows.

### 4.3 Trends and Insights

Figures 5a and 5b indicate a clear trend towards the increased adoption of non-interpretable models over time. Initially, both interpretable and non-interpretable models were employed as baseline models; however, by 2023, non-interpretable models have dominated as the winning models. In particular, ensemble and deep learning models have become the most common categories, especially in 2023. This trend highlights the healthcare sector’s growing reliance on complex models that, while less transparent, offer superior predictive accuracy.

**FIGURE 5.**
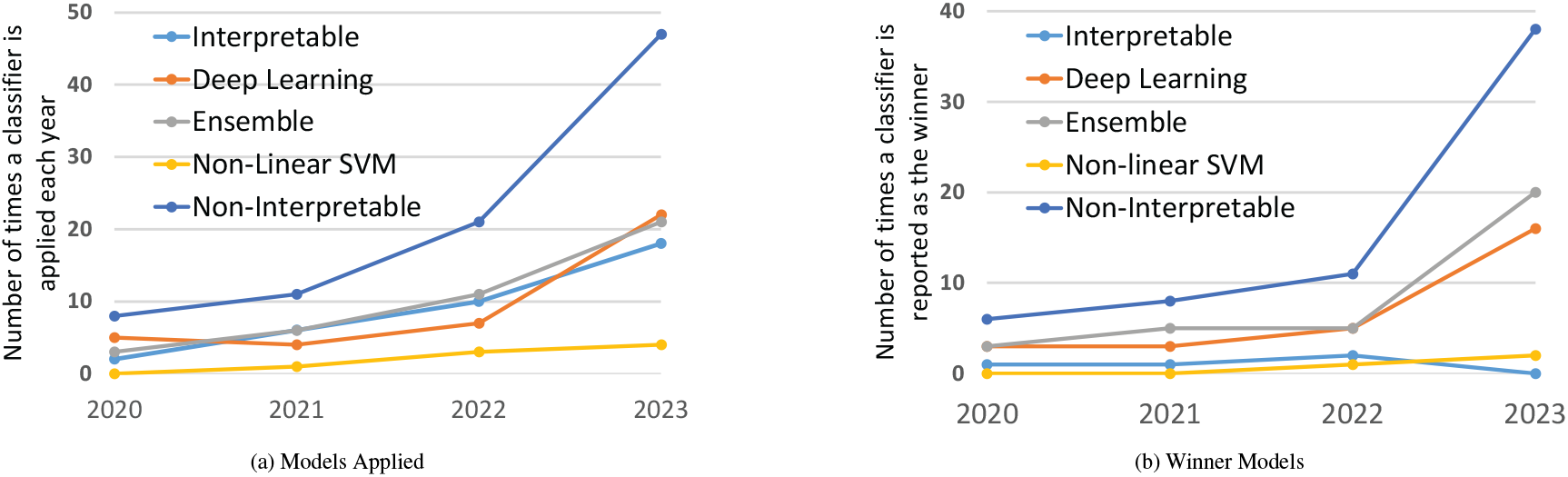
Baseline ML models and winners’ trends from Jan 2020 to Dec 2023 are shown; data for 2024 is not shown as it is an ongoing year.

**FIGURE 6.**
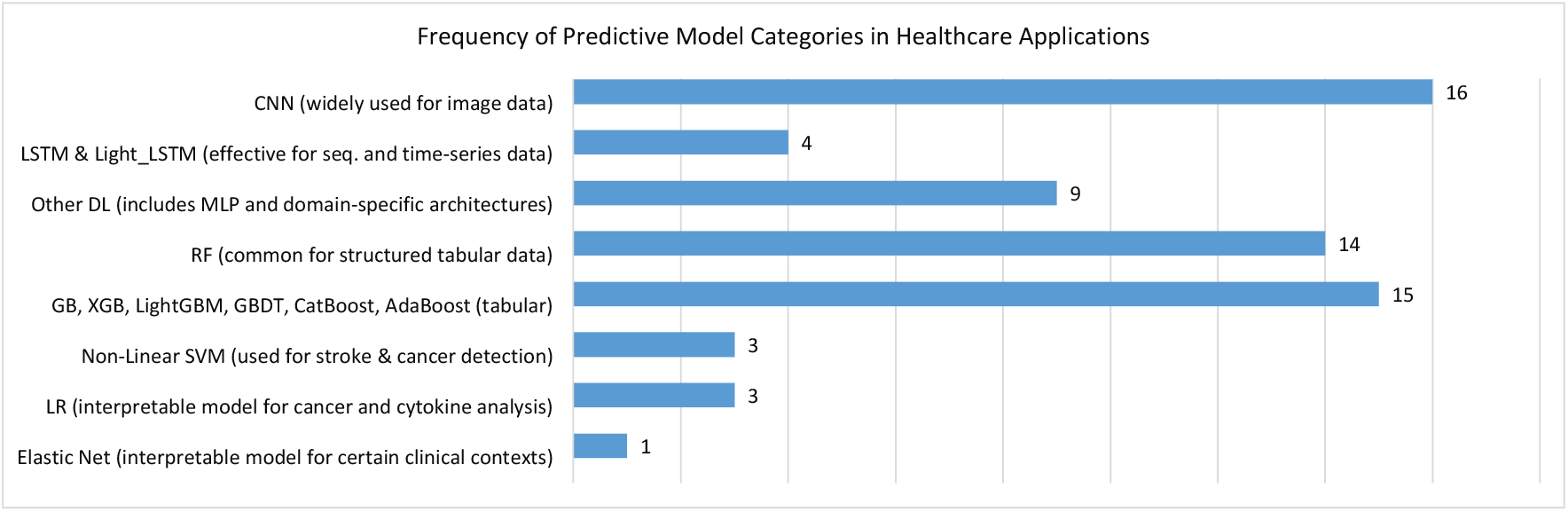
Frequency of Predictive Model Usage in Healthcare Applications.

Table 6 lists the categories of models and their frequency count as best-performing, revealing a varied range of ML models, each offering unique advantages:

**TABLE 6.**
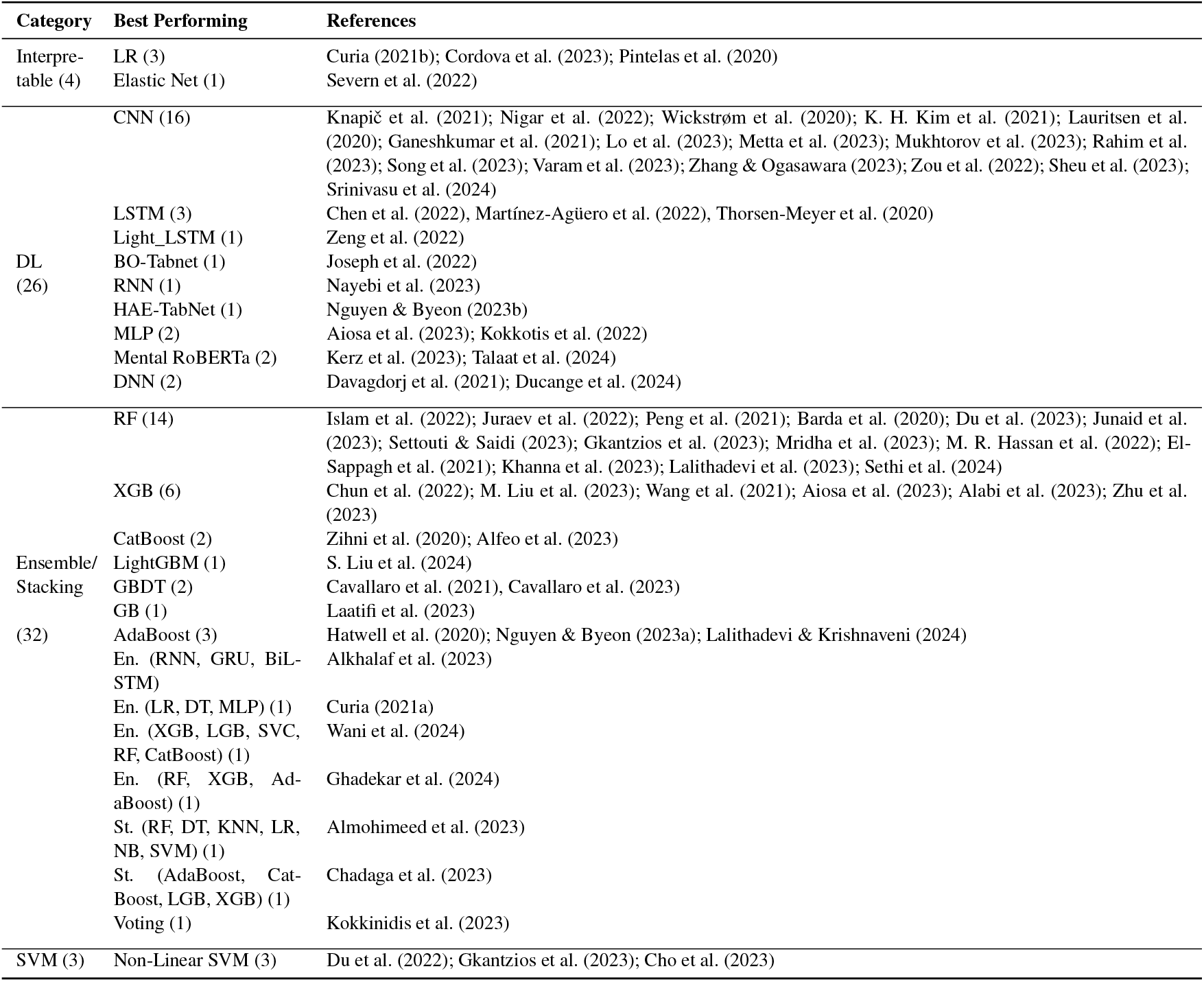
Categorisation of the best-performing ML models.

- **Convolutional Neural Networks (CNNs):** Ideal for image data in skin lesion classification, endoscopy, brain diseases, cancer, viruses, and cardiovascular conditions.
- **LSTM**: Effective for time-series and sequential data (Martínez-Agüero et al., 2022; Thorsen-Meyer et al., 2020).
- **Random Forest (RF) and Ensembles:** RF demonstrates high performance in structured tabular medical data, including predictions related to liver disease, diabetes, cardiovascular conditions, and COVID-19. Furthermore, ensembles are robust due to their ability to aggregate predictions from multiple models, performing well across various medical domains, including image and tabular data (Alkhalaf et al., 2023; Curia, 2021a; Debjit et al., 2022; Wani et al., 2024).
- **Support Vector Machines (SVMs):** Effective for medical predictions, particularly in gestational diabetes (Du et al., 2022), stroke outcomes (Gkantzios et al., 2023), and cancer detection (M. R. Hassan et al., 2022).
- **Interpretable Models:** Logistic regression and Elastic NET have shown noteworthy performance in specific clinical contexts, such as cancer (Curia, 2021b; Cordova et al., 2023; Pintelas et al., 2020) and cytokine analysis (Curia, 2021b).

### 4.4 Challenges in Model Implementation

Implementing predictive models in clinical settings presents challenges, particularly concerning data heterogeneity, model interpretability, and the integration of XAI methods. While tree-based models facilitate feature importance analyses, neural networks require complex, post-hoc explanations. The variability in data types across application areas necessitates careful model selection, highlighting the need for tailored approaches and comprehensive comparative studies.

The analysis reveals several key gaps in the current use of predictive models in healthcare. A significant challenge is the increasing reliance on non-interpretable models, such as deep learning and ensemble methods, which, despite their superior predictive performance, lack transparency and hinder their adoption in clinical settings. This gap underscores the need to develop more interpretable models or effective XAI techniques tailored to these complex models. Furthermore, comparative studies are scarce in evaluating the effectiveness of different models and XAI methods across various healthcare domains. This lack of comprehensive evaluation limits our understanding of the trade-offs between model complexity, performance, and interpretability. The application-specific selection of models suggests a need for hybrid approaches that can leverage the strengths of different models to address the unique challenges presented by diverse healthcare data.

Additionally, the inherent heterogeneity of healthcare data across different contexts highlights the necessity for developing more robust models that can generalise across varied data types, ensuring reliable and consistent performance. Addressing these gaps is critical for advancing the integration of AI in healthcare and ensuring that predictive models can be both powerful and clinically relevant.

## 5 XAI METHODS

This section explores the various XAI methods used in the reviewed studies, categorising them based on their applicability, approach, and relevance to different types of machine learning models. The selection of XAI methods is crucial for interpreting complex models, especially in healthcare, where transparency and trust are vital. Readers seeking a descriptive understanding of popular XAI methods discussed can refer to the table in the Appendix (Table 4), which provides detailed descriptions and healthcare applications of these methods, complementing the main discussion.

Figure 7 presents the frequency of various XAI methods used across healthcare studies. This visualization provides an overview of the adoption of different techniques, highlighting SHAP and LIME as the most widely used methods, followed by GradCAM and its variants. Such a depiction aids in understanding the trends and preferences in selecting XAI methods for healthcare applications.

**FIGURE 7.**
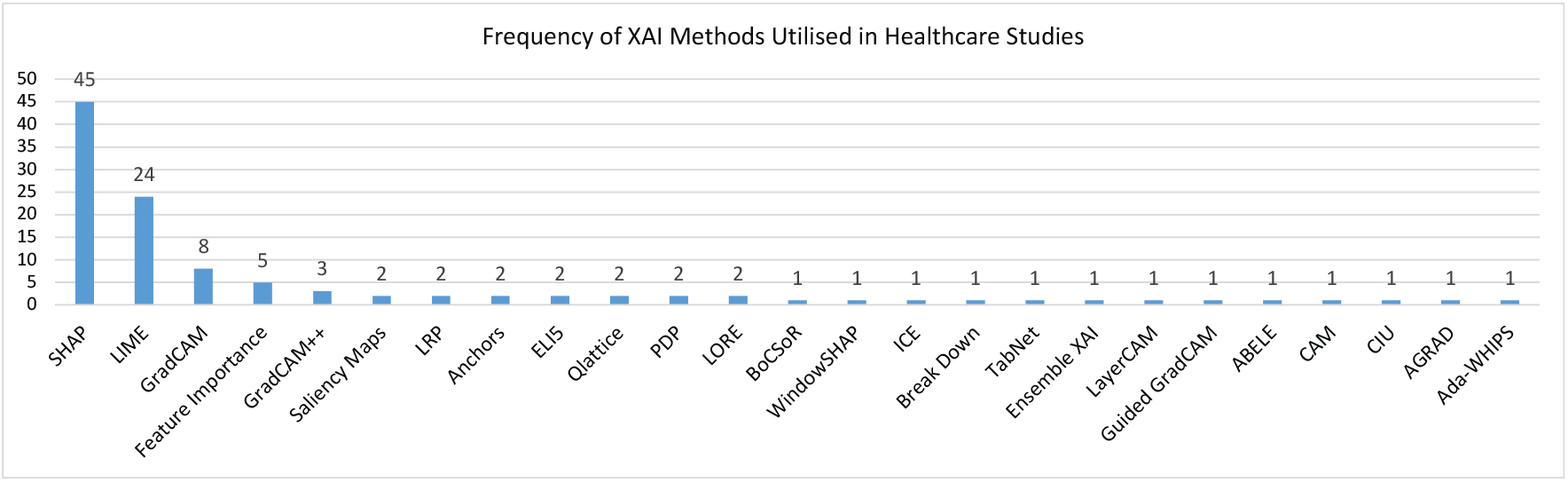
Frequency of XAI Methods in Healthcare Research.

The section begins with a framework for categorising XAI methods, introducing five key dimensions to categorise the XAI methods used in healthcare studies. Following this, the selection of XAI methods based on model types subsection discusses how the choice of XAI methods varies according to the types of ML models used. The effectiveness and challenges of XAI methods subsection then examines the strengths and limitations of these methods in terms of interpretability, computational complexity, and applicability across different data types. Finally, the section concludes with open challenges, which identifies the remaining barriers to effective integration of XAI in clinical settings and suggests future directions for research and development.

### 5.1 Framework for Categorising XAI Methods

XAI methods can be defined based on five key aspects or dimensions: *stage, applicability, scope, form*, and *type* (Schwalbe & Finzel, 2023). These dimensions offer a comprehensive framework for understanding how and when different XAI methods can be effectively employed in different contexts.

The *stage* dimension refers to the timing of the explanation generation, distinguishing between post hoc and ante hoc methods. Post hoc methods generate explanations after the model has been constructed, while ante hoc methods involve models designed to be interpretable from the outset. For instance, SHAP is a post hoc method that calculates Shapley values to fairly assign feature importance for model predictions, widely used in healthcare for interpreting complex models in tasks like patient risk stratification and diagnosis (Chen et al., 2022; Junaid et al., 2023; Severn et al., 2022; Ducange et al., 2024; Talaat et al., 2024). GradCAM, another post hoc method, generates localisation maps to highlight critical regions in medical images, such as aiding in disease diagnosis (K. H. Kim et al., 2021; Lo et al., 2023; Zhang & Ogasawara, 2023). In contrast, TabNet is an ante hoc method designed for tabular data, employing sequential attention to ensure interpretability and accuracy like those involving electronic health records (Joseph et al., 2022).

The *applicability* of XAI methods can be either model-agnostic or model-specific. Model-agnostic methods, such as LIME and SHAP, can be applied across various ML models. LIME, for instance, generates a new dataset of permuted samples and trains an interpretable model to explain individual predictions, making it valuable in personalised medicine where patient-specific outcomes are critical (Junaid et al., 2023; Curia, 2021a; Talaat et al., 2024). In contrast, model-specific methods are tailored to particular models, like GradCAM, designed for deep learning models such as CNNs. GradCAM++ extends this by offering more detailed class activation maps, particularly useful in complex medical imaging tasks where multiple objects are present (Mukhtorov et al., 2023; Zou et al., 2022).

The *scope* of XAI methods can be global or local. Global methods, such as PDP, explain the overall behaviour of a model by showing the effect of individual features on predictions while holding other features constant. In healthcare, PDPs illustrate how factors like blood pressure or cholesterol levels impact disease risk (Chen et al., 2022; Peng et al., 2021). Conversely, local methods focus on individual predictions; for example, Anchors use high-precision rules to clarify specific decisions made by classifiers, which is crucial for providing explanations in complex clinical decision support systems (Khanna et al., 2023; Hatwell et al., 2020).

The *form* of the explanation encompasses both rule-based and visual representations. Rule-based explanations, exemplified by Anchors, provide clear conditions that lead to specific predictions, offering straightforward interpretability essential for clinicians to trust the decision-making process. In contrast, visual representations like Saliency Maps and GradCAM emphasise regions of input data that significantly influence predictions, making them particularly effective in medical imaging for identifying key areas, such as tumorous regions in scans.

XAI methods output different *types* of explanations, including feature importance rankings and visualisations. Feature importance rankings, provided by methods like SHAP, reveal the influence of individual features on predictions. Visualisations, such as PDP and ICE plots, demonstrate the impact of feature changes on predictions, with ICE plots providing insights for personalised medicine by showing how feature variations influence individual outcomes (Chun et al., 2022).

Table 7 provides an overview of the XAI methods utilised in the literature, detailing their five aspects or dimensions and the types of ML models they were applied to. It is evident from the table that SHAP is the most widely used explainer method applied across various categories of ML models. Following SHAP, LIME is the second most popular choice, with both being model-agnostic methods. GradCAM is preferred for deep learning models among model-specific methods, particularly for image-based tasks.

**TABLE 7:**
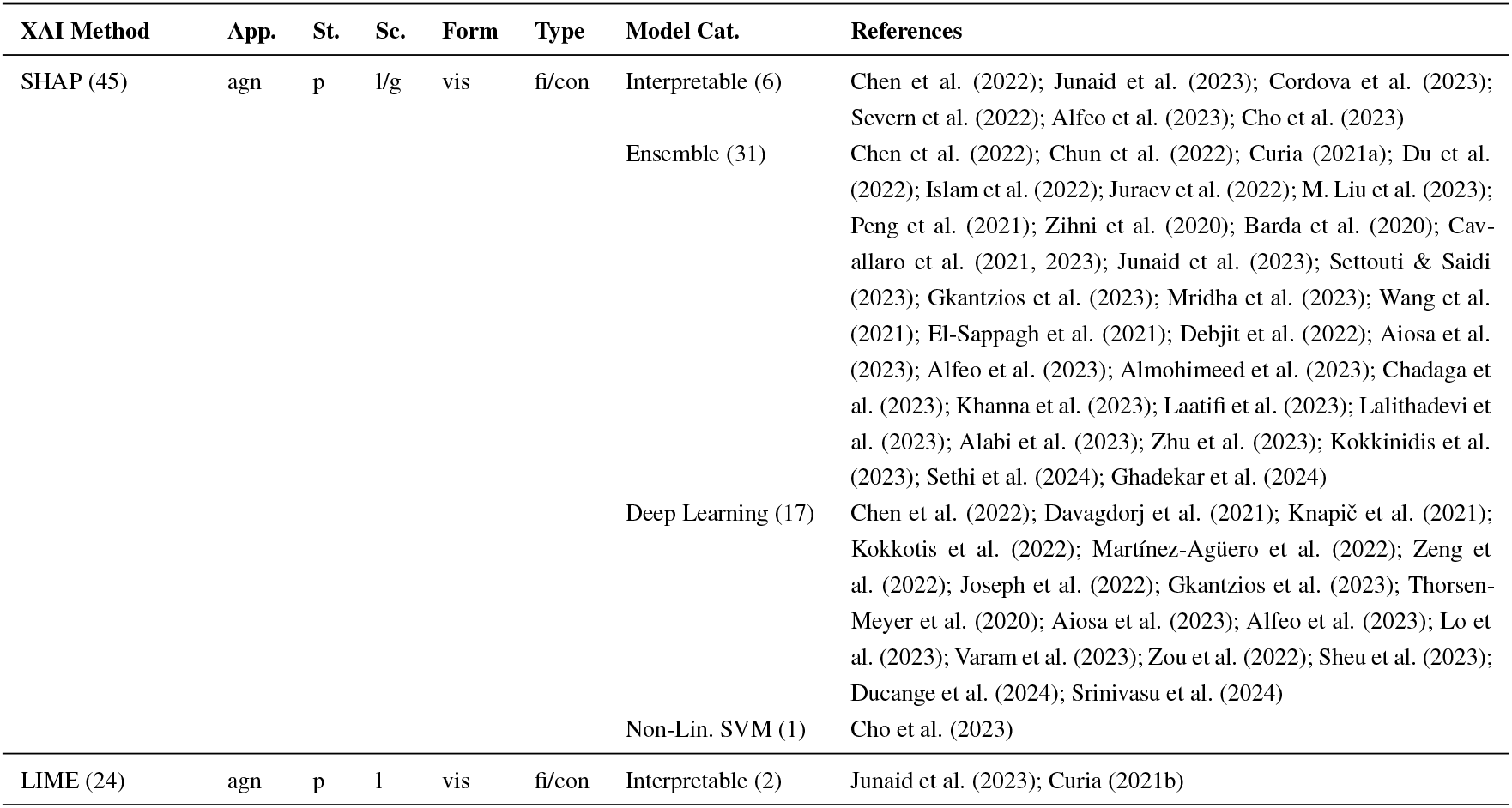
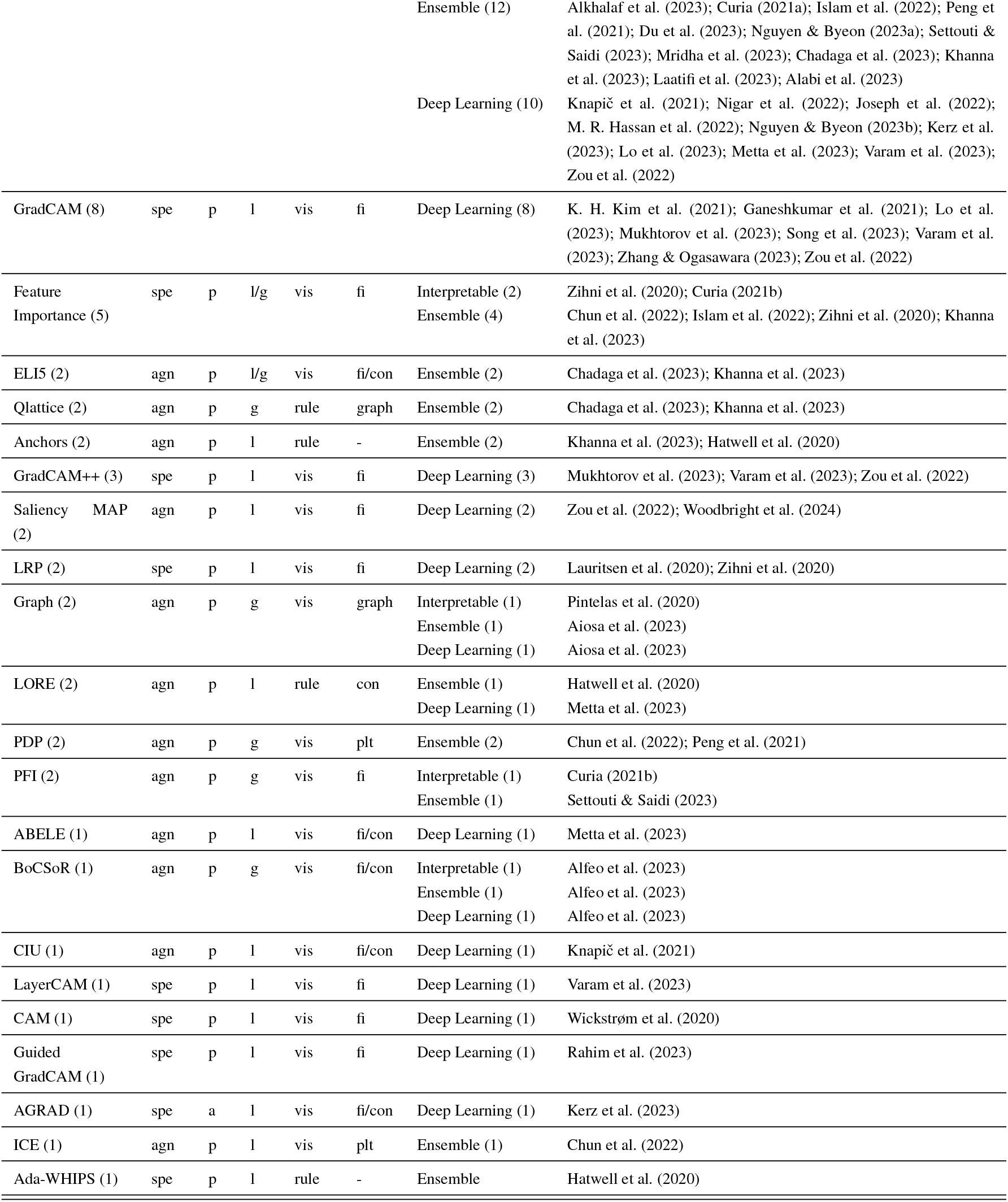
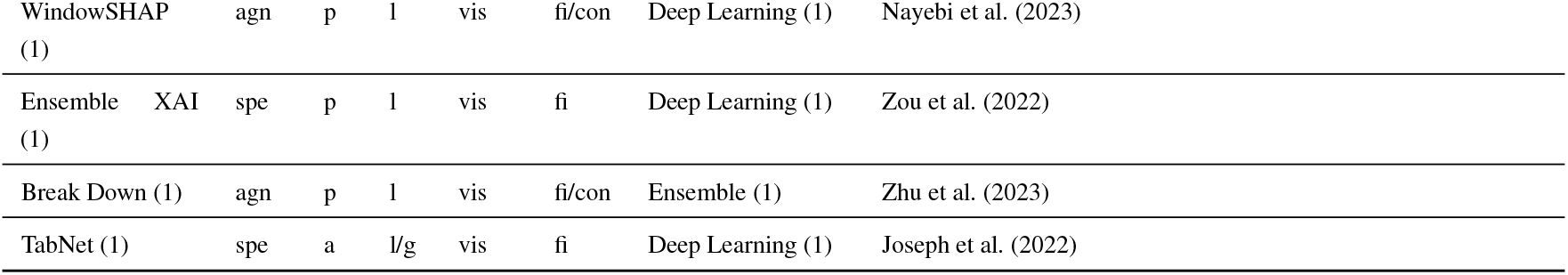
Methods for Explainability. Abbreviations by column *Applicability*=App., Agnostic=agn, Specific=spe; *Stage*=St., post hoc=p, ante hoc=a; *Scope*=Sc., local=l, global=l; visual=vis; feature importance=fi, contrastive=con, plot=plt.

Regarding the *stage*, most methods are post hoc, except AGRAD and TabNet, which are ante hoc methods utilised in conjunction with deep learning and attention-based mechanisms. Regarding the *scope*, there is a strong emphasis on local methods, reflecting the importance of explaining individual instances or patient records over the broader behaviour of a condition, class, or disease, although global scope methods are also utilised. Visualisation is the preferred output format among the selected methods, providing a straightforward means of explaining decisions, followed by rule-based approaches that explain reasoning through feature constraints. The output type is determined mainly by the importance of features in influencing a decision, typically involving comparing each feature’s impact on an automated decision (such as identifying correlated or influential features in a specific decision). Additionally, there are occasional instances of graphs and plots that explain the decision-making process.

### 5.2 Selection of XAI Methods Based on Model Types

The choice of XAI methods is often guided by the predictive model and the specific requirements of the healthcare application:

- **Neural Networks**: Deep learning models, particularly CNNs for image classification, often utilise GradCAM and its variants (e.g., GradCAM++, Guided GradCAM) to provide visual explanations by highlighting important regions in images. This is crucial for diagnosing conditions in medical imaging (K. H. Kim et al., 2021; Lo et al., 2023; Zhang & Ogasawara, 2023). Additionally, Saliency Maps help clarify how different input components affect the final output, enhancing the understanding of complex model behavior (Chen et al., 2022; Junaid et al., 2023).
- **Tree-Based Models**: For models such as RFs and GBMs, feature importance methods are commonly utilised. These methods identify the most influential features in the model’s predictions, thereby providing insights into the decision-making process (Curia, 2021a; Junaid et al., 2023). PDPs are also employed to visualise the relationship between individual features and the predicted outcome while keeping other features constant, helping to understand how specific factors like blood pressure impact disease risk Peng et al. (2021). SHAP has also been applied to various tree-based ensembles to enhance understanding of decision-making (Sethi et al., 2024; Ghadekar et al., 2024).
- **Support Vector Machines (SVMs)**: SVMs often use model-agnostic methods like LIME or SHAP for post-hoc explanations due to their less interpretable nature. LIME generates local explanations by approximating the decision boundary of an SVM with a simpler model, while SHAP provides detailed feature importance by calculating Shapley values (Junaid et al., 2023; Cho et al., 2023).
- **Logistic Regression and Other Linear Models**: Despite their inherent interpretability due to their linear structure, methods like SHAP can offer more granular insights into feature interactions, especially in complex scenarios where understanding the subtle impact of features is crucial (Severn et al., 2022).

### 5.3 Effectiveness and Challenges of XAI Methods

While XAI methods enhance model interpretability, they face several challenges:

- **Interpretability vs. Fidelity**: There is often a trade-off between the simplicity of the explanation and its accuracy in reflecting the models true decision-making process. For instance, LIME offers easy-to-understand explanations but cannot truly capture the global behaviour of complex models (Chun et al., 2022).
- **Computational Complexity**: Some XAI methods, particularly model-agnostic ones like SHAP, are computationally intensive, making them less suitable for real-time applications. This is a significant consideration in healthcare, where timely decisions are critical (Severn et al., 2022; Cho et al., 2023).
- **Applicability Across Different Data Types**: The effectiveness of XAI methods varies with data type. For instance, Grad-CAM excels with image data but is not applicable to tabular data, whereas methods like SHAP and feature importance are more versatile across different data types (K. H. Kim et al., 2021; Junaid et al., 2023).
- **Model-Specific Constraints**: Model-specific XAI methods are limited by the architecture for which they are designed, restricting their use in diverse scenarios. This necessitates the development of more adaptable tools that can be used across various model types and applications (Curia, 2021a; Cho et al., 2023).

### 5.4 Open Challenges for XAI in Healthcare

Ongoing research in XAI should prioritise the balance between interpretability and computational efficiency to facilitate realtime clinical decision-making (Severn et al., 2022; Cho et al., 2023). A hybrid approach that combines model-agnostic and model-specific methods can offer more comprehensive and context-sensitive explanations by merging visual and rule-based techniques (K. H. Kim et al., 2021; Junaid et al., 2023). Moreover, a strong emphasis on user-centric design is crucial to developing intuitive, user-friendly tools that integrate seamlessly into clinical workflows (Curia, 2021a; Cho et al., 2023). These advancements will significantly enhance the practical utility and effectiveness of XAI in healthcare, thereby promoting greater transparency and trust in clinical decision-making.

## 6 STUDIES UTILISING MULTIPLE XAI METHODS

This section provides an analysis of twenty-five studies that employ multiple XAI methods to enhance model interpretability in healthcare applications. It begins by exploring comparative studies that directly evaluate the effectiveness of different XAI methods across various data types, including image and tabular data. To support this analysis, a stacked bar has been included as Figure 8. This visualisation highlights the combinations of methods applied in various domains, with Neurology and Cancer being the most frequently explored areas. The next subsection highlights the complementary use of multiple XAI methods, demonstrating their combined utility in diverse clinical scenarios, although without a formal comparison. Finally, the section concludes with an examination of open issues and future directions, discussing the challenges and potential solutions for effectively integrating multiple XAI methods in healthcare settings.

**FIGURE 8.**
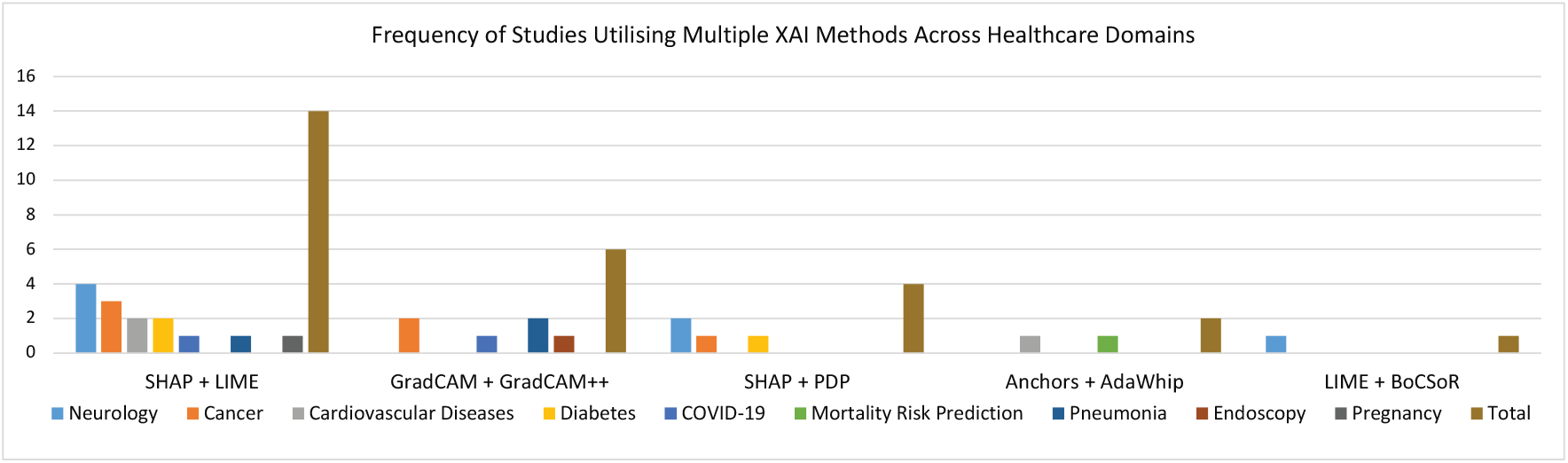
This stacked bar chart represents the number of studies employing combinations of XAI methods in various healthcare domains. It highlights the prevalence of methods such as SHAP + LIME and GradCAM + GradCAM++ in Neurology and Cancer.

### 6.1 Comparative Studies on Multiple XAI Methods

Ten studies directly compared various XAI methods to explain prediction outcomes in healthcare. Six of these studies focused on image data, all using deep learning techniques and employing three to six XAI methods. The dominant comparison strategies included heatmaps, user studies, saliency maps, and explainability scores. GradCAM and its variants were the most frequently used methods, followed by LIME, SHAP, CIU, and ABELE, each providing distinct strengths in different contexts. In contrast, studies focusing on tabular data primarily utilised ensemble methods, sometimes incorporating deep learning and non-linear SVMs. These studies typically employed two to three XAI methods, with particular emphasis on methods like AdaWhip and BoCSoR, favoured for their performance and complementary roles.

Table 8 provides a concise overview of studies that utilised multiple XAI methods and compared them. For image data:

**TABLE 8.**
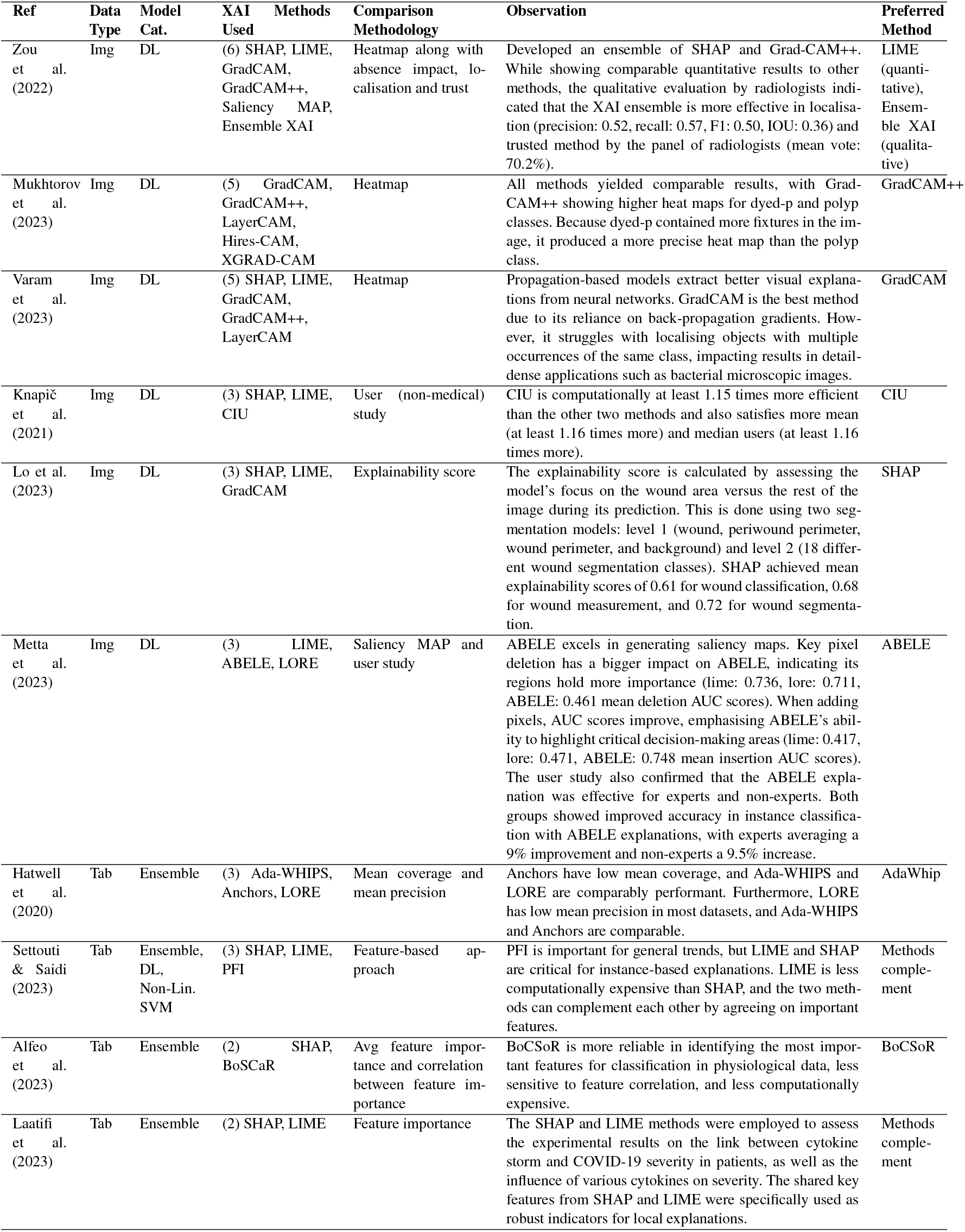
Summary of studies that used multiple XAI methods and compared them.

- In a study by Zou et al. (2022), LIME achieved better quantitative scores, but radiologists preferred the Ensemble XAI method for its effectiveness in localisation and trust, shown through heatmap evaluations.
- GradCAM++ was identified as the top-performing method for endoscopic analysis in Mukhtorov et al. (2023), based on heatmap evaluation, while it excelled in visual explanations during backpropagation, as noted in Varam et al. (2023). SHAP and LIME were also acknowledged for their efficacy in feature-based explanations.
- LIME, SHAP, and CIU were evaluated based on human comprehension, satisfaction scores, computational complexity, and overall understanding in Knapič et al. (2021). CIU was highlighted as the superior method compared to LIME and SHAP.
- SHAP was preferred over LIME and GradCAM for explaining decisions, based on an *explainability score* in Lo et al. (2023), which measured the model’s focus on the wound area versus the rest of the image.
- ABELE was found to be more effective than local explainers LIME and LORE in skin lesion detection, as shown in Metta et al. (2023), due to its superior saliency maps despite higher computational complexity.

For tabular data:

- Hatwell et al. (2020) compared Anchors, AdaWhip, and LORE based on mean coverage and mean precision, with AdaWhip outperforming the other two.
- Settouti & Saidi (2023) evaluated SHAP, LIME, and permutation feature importance (PFI) based on feature importance values and computational complexity. PFI emerged as the most efficient method for general understanding. At the same time, LIME and SHAP proved valuable for individual instances, with consensus among methods underscoring the significance of distinctive features for each cancer patient.
- A novel XAI method, BoCSoR, outperformed SHAP in feature correlation values for social and emotional tasks, as demonstrated in a study by Alfeo et al. (2023).
- Laatifi et al. (2023) used LIME and SHAP to identify robust indicators of COVID-19 severity. The key features identified by both methods were deemed robust indicators for each patient’s record.

### 6.2 Complementary Use of Multiple XAI Methods

17 additional studies employed multiple XAI methods without direct comparison but highlighted their complementary usefulness. Fourteen of these studies focused on tabular data, with one involving text data. These studies confirmed the validity of XAI methods by aligning the identified important features with previous research, demonstrating the potential of XAI in unexplored domains such as early detection of Parkinsons disease (Junaid et al., 2023), assessing cancer risk (Curia, 2021a), and supporting healthcare professionals in various clinical scenarios (Aiosa et al., 2023). The results are outlined in Table 9, with the following key characteristics of these studies:

**TABLE 9.** Summary of studies that used multiple XAI methods but did not compare them.

| Ref | Data Type | Model Cat. | XAI Methods Used | Observation |
| --- | --- | --- | --- | --- |
| Khanna et al. (2023) | Tab | Ensemble | (6) SHAP, LIME, FI, ELI5, Qlattice, Anchor | The XAI methods have been validated by identifying the important features, which were confirmed by previous research. The research discovered that anomalies in features such as respiratory rate, blood pressure, body temperature, calcium, and lactate levels positively contribute to patient severity. |
| Chadaga et al. (2023) | Tab | Ensemble | (4) SHAP, LIME, ELI5, Qlattice | XAI methods were used to find important markers for screening COVID-19 patients. The important markers found were albumin, ALT, basophil, and TWBC. The authors said that XAI methods can help healthcare professionals in situations like COVID-19 when the best solution is unknown and be helpful in the first screening of coronavirus patients. |
| Chun et al. (2022) | Tab | Ensemble | (4) SHAP, ICE, FI, PDP | FI and PDP helped scrutinize the most important global features concerning cognitive impairment, with ICE and SHAP allowing the interpretation of specific patient data. Furthermore, using graphs, patients can better understand the neuropsychological factors at risk, which is a step towards precision medicine. Key discoveries from the explanations include RCFT delayed recall, CDR-SOB, age, K-MMSE, COWAT-animal, education, SVLT delayed recall, RCFT copy time, and APOE genotype - all of which resonate with previous research. |
| Peng et al. (2021) | Tab | Ensemble | (3) SHAP, LIME, PDP | PDP and SHAP (via mean SHAP values) were used as global explainers, and LIME as a local explainer. These methods found that ascites, spiders, bilirubin, albumin, malaise, varices, and the SpleenPalpable feature had more impact than the others, which aligns with prior knowledge from hepatobiliary physicians. |
| Curia (2021b) | Tab | Interpretable, Ensemble | (3) LIME, FI, PFI | In a stacked-based clustering approach, permutation importance was used to evaluate the significance of each clustering method in grouping a set of features to identify distinct patterns for accurate classification. Feature importance served as the global method, while LIME was employed as the local explainer. The features identified through explainers confirmed the results obtained and supported previous studies. |
| Islam et al. (2022) | Tab | Ensemble | (3) LIME, SHAP, FI | SHAP was used to assess global feature importance, and LIME was used as a local explainer. The feature "had_previous_c_section" was identified as one of the most important global features, and LIME identified contributing features, including "suffered_domestic_violence." XAI methods were argued to provide an early explainable predictive approach to assist in implementing new policies to reduce unnecessary C-Section deliveries. |
| Joseph et al. (2022) | Tab | DL | (3) SHAP, LIME, TabNet | Uses SHAP and TabNet as global explainers and LIME as a local explainer. The collective inference suggests that insulin and polyuria are significant features associated with diabetes risk. TabNet, an ante-hoc method, also demonstrates high accuracies on various datasets. |
| Zihni et al. (2020) | Tab | Ensemble, Interpretable, DL | (3) SHAP, FI, LRP | Took a feature-based approach and used Shapley values for ensembles, model coefficients for logistic regression and deep Taylor decomposition for deep learning to explain respective models. |
| Alabi et al. (2023) | Tab | Ensemble | (2) SHAP, LIME | Globally, SHAP showed that age, T-stage, ethnicity, M-stage, marital status, and grade were key factors for NPC patient survival. LIME and SHAP methods demonstrated how each feature impacted individual predictions, confirming globally important features. |
| Mridha et al. (2023) | Tab | Ensemble | (2) SHAP, LIME | SHAP as a global explainer identified Age, Avg. Glucose Level, Work Type, Residence Type, Gender, and Ever Married as important features. This generally aligns with the views of hepato-biliary experts and previous research. Lime was used to reveal local features for individual patients, highlighting instances where Age and Work Type were occasionally ranked higher than other features for stroke patients, echoing observations in globally important features. |
| Junaid et al. (2023) | Tab | Ensemble | (2) SHAP, LIME | Used SHAP for global and local explanations and LIME for local explanations to detect Parkinson's disease early. The research compared the outputs of SHAP and LIME for RF and LightGBM models. XAI methods highlighted the NP3BRADY feature as the most important, while local explainers identified the MESEADLG feature as the best feature for both models. |
| Curia (2021a) | Tab | Ensemble | (2) SHAP, LIME | LIME and SHAP were used as local explainers. It was suggested that providing explanations for decision-making to medical professionals in scenarios like predicting cancer risk serves the purpose, even if methods share some features and differ in others. Severity perception was most indicative of cervical cancer in two cases, confirming a general understanding. |
| Aiosa et al. (2023) | Tab | Ensemble, DL | (2) SHAP, Graph | The study examined the links between obesity, diabetes, cardiovascular issues, and heart disease. It utilised SHAP to provide insights globally and locally. Local findings were showcased through three case studies featuring patients with positive test results for multiple diseases, negative results, and varied predictions for comorbidities. A multi-node graph was used to aid healthcare professionals and patients in understanding the progression of these conditions. The approach assists clinicians in forecasting pathologies. |
| Zhu et al. (2023) | Tab | Ensemble | (2) SHAP, Break Down | Uses SHAP for global and Break Down as local explainer. Overall, insights. 1- clinical significance is highlighted through top features. 2- higher values of features, such as myocardial enzyme spectrum markers and diabetes-associated markers, lead to BD, while lower values contribute to MDD. 3- potential for discovery. 4- offers general recommendations. |
| Kerz et al. (2023) | Text | DL | (2) LIME, AGRAD | Uses LIME and AGRAD to identify the word categories relied on by models when making predictions. Explainers reveal that multi-task fusion models learned significant correlations between mental health conditions, emotions, and personality traits. |

- Studies by Khanna et al. (2023); Chun et al. (2022); Peng et al. (2021); Curia (2021b); Mridha et al. (2023); Zhu et al. (2023) validated XAI methods by confirming important features through previous research.
- Some studies showed that XAI methods were instrumental in identifying crucial features related to medical conditions for which definitive solutions are yet to be discovered, as seen during the COVID-19 pandemic (Chadaga et al., 2023). They can also be instrumental in unexplored research domains like the early detection of Parkinsons disease (Junaid et al., 2023), assessing cancer risk (Curia, 2021a), cancer survival Alabi et al. (2023), supporting healthcare professionals and patients (Aiosa et al., 2023), and addressing mental health conditions (Kerz et al., 2023; Zhu et al., 2023).
- Islam et al. (2022) proposed XAI methods as an early predictive approach to aid in implementing policies to reduce unnecessary C-Section deliveries.
- Joseph et al. (2022) demonstrated the effectiveness of collective inference using ante hoc and posthoc XAI methods, and in a study by Zihni et al. (2020), various XAI methods were employed to explain distinct ML models.

### 6.3 Open Issues and Future Directions

While using multiple XAI methods enhances the interpretability of AI models in healthcare, several challenges remain. Integrating different methods often lacks standardisation, leading to inconsistent or conflicting interpretations. Computational efficiency is also a concern, as combining multiple XAI techniques can increase the complexity and cost of real-time analysis. Moreover, there is a risk of information overload for end users, such as clinicians, which can impact trust and acceptance. Addressing these challenges requires developing standardised frameworks for integrating XAI methods, optimising computational efficiency, and designing user-centric explanations that balance detail and clarity. Establishing comparative evaluation standards (i.e., XAI metrics like fidelity and faithfulness, as well as user studies, which are currently overlooked) will also be crucial for identifying the most effective methods for various healthcare applications. Future research should focus on overcoming these open issues to fully leverage multiple XAI methods, ensuring they are robust, efficient, and user-friendly for clinical use.

## 7 OPEN RESEARCH GAPS AND RECOMMENDATIONS

Despite advancements in XAI methods within healthcare, several critical research gaps need to be addressed to improve the effectiveness and adoption of these technologies. Each identified gap is paired with a recommendation to bridge the gap and advance the field.

### 1. Limited Public Datasets and Uneven Application Coverage

- **Gap:** Over 60% of the datasets used in the reviewed studies are private (Chun et al., 2022; Juraev et al., 2022; Martínez-Agüero et al., 2022), particularly in critical areas like neurological disorders (only 6 out of 19 datasets are public), ICU mortality risk (all private), and antimicrobial resistance (all private). While areas like neurological conditions and cancer are well-researched, others, such as antimicrobial resistance and hepatitis, remain underexplored (Martínez-Agüero et al., 2022; Peng et al., 2021).
- **Recommendation:** Prioritise open access to high-quality medical datasets by anonymising patient data, obtaining patient consent, and enforcing data governance practices. This will enhance transparency and accessibility for XAI research in healthcare, facilitating the development of replicable models and enabling comparative studies that are currently hindered by the scarcity of public datasets. Furthermore, encouraging research in underexplored healthcare areas and creating public datasets will promote a more balanced investigation of XAI applications, advancing the field.

### 2. Ineffective Data Treatment Methods

- **Gap:** Many tabular datasets contain missing values, noise, and outliers. Current strategies, such as simple imputation or dropping data with significant missing values, often prove inadequate, especially in small datasets (Zhu et al., 2023). Also, in multiple datasets, data is unevenly distributed among different classes, leading to models biased toward majority classes (Chun et al., 2022; Junaid et al., 2023; Settouti & Saidi, 2023; Lauritsen et al., 2020) and performing poorly for minority classes. There is also limited application of targeted feature engineering.
- **Recommendation:** Develop advanced, especially “medically informed” data preprocessing techniques to address missing values, noisy data, and outliers. For data balancing challenges, explore methods like SMOTE, GANs, counterfactual data augmentation (Qureshi et al., 2024), and ensemble approaches, all while considering medical constraints. These strategies can reduce biases and enhance model performance, especially for minority classes. Also, increase the use of feature selection and engineering methods tailored to medical contexts to optimise clinically relevant features, enhancing model accuracy and benefitting interpretability by design.

### 3. Data Diversity and Multimodal Integration

- **Gap:** Many studies focus mainly on tabular data, often overlooking the integration of other data types, such as imaging or genomic data (Hatwell et al., 2020; Joseph et al., 2022).
- **Recommendation:** XAI studies should incorporate diverse data types to enhance model generalisability and robustness. Using multimodal data (Cremonesi et al., 2023; S. Y. Kim et al., 2024) including imaging, genomic, and tabular data, can provide a deeper understanding of various health conditions and their complex, multifactorial nature, which is crucial for capturing intricate relationships in healthcare.

### 4. Broader Use of XAI Methods and Reliable Solutions

- **Gap:** Many studies focus on single XAI methods (Alkhalaf et al., 2023; Ganeshkumar et al., 2021), which may limit a comprehensive understanding of interpretability. There is also a pressing need for reliable XAI solutions that provide clear, understandable explanations, particularly in light of ethical and regulatory concerns.
- **Recommendation:** Encourage using multiple XAI methods (Zou et al., 2022; Khanna et al., 2023) within studies to gain a more complete and robust understanding of model interpretability, leveraging the complementary strengths (Joseph et al., 2022) of different approaches. Additionally, develop explainability-focused model designs that adhere to regulatory and ethical standards (Lyle et al., 2023; Jung et al., 2024; Wabro et al., 2024) to enhance trust in AI-driven healthcare decisions.

### 5. Lack of Comparative Evaluation Using XAI Metrics

- **Gap:** There is a significant gap in comparative studies evaluating different XAI methods using diverse XAI metrics (see Table 8). Also, there is no consensus on standardised evaluation metrics for XAI, making it difficult to compare the effectiveness of different methods (see Table 9).
- **Recommendation:** Future research should focus on comparative studies assessing various XAI methods with XAI metrics, such as fidelity, faithfulness, interpretability, clarity, completeness, stability, and relevance (Pawlicki et al., 2024; Kostopoulos et al., 2024). These should examine model-agnostic versus model-specific approaches and output types in different medical contexts. In radiology, fidelity is crucial to align explanations with imaging data, while interpretability is vital in primary care for clinician trust. The evolving practice of standardised XAI metrics in healthcare will enhance formal study comparisons and aid in addressing ethical challenges.

### 6. Challenges in Real-World Integration and Cybersecurity

- **Gap:** Integrating XAI methods into clinical workflows presents practical challenges, including computational demands, compatibility with existing healthcare systems, cybersecurity concerns, and specialised training for healthcare professionals.
- **Recommendation:** Design XAI systems that can be seamlessly integrated into clinical environments, addressing computational requirements Pumplun et al. (2023) and ensuring robust cybersecurity measures Rjoub et al. (2023) to protect patient data. This will require collaboration Nasarian et al. (2024) among AI researchers, healthcare professionals, and cybersecurity experts to develop secure and reliable XAI systems.

## 8 CONCLUSION

This article provides a comprehensive review of Explainable AI (XAI) in healthcare, focusing on advancements in methodologies and the challenges hindering their practical implementation. The increasing adoption of complex models, such as deep learning and ensemble methods, particularly in areas like medical imaging, cardiovascular disease prediction, and neurological disorder diagnosis, has heightened the need for robust XAI techniques. Ensuring the interpretability of these models remains a critical challenge that must be addressed to foster transparency and trustworthiness in clinical environments.

Key obstacles identified include the scarcity of publicly available datasets, particularly in neurological disorders and ICU mortality risk, which limits replicability and comparative studies. Data imbalances, inadequate preprocessing, and underutilisation of feature selection and engineering techniques further exacerbate the challenges in achieving model fairness and interpretability. Addressing these gaps is crucial to improving the accuracy and effectiveness of AI models in healthcare decision-making.

To advance the field, there is an urgent need for explainability-centric models that prioritise user-friendly insights without compromising predictive performance. Comparative studies using standardised XAI metrics are essential for establishing best practices and ensuring consistent assessments. Leveraging multiple XAI methods within studies can offer a more comprehensive understanding of model behaviour. Overcoming practical challenges, such as computational demands and the integration of XAI into clinical workflows, will require interdisciplinary collaboration. By addressing these issues, XAI can play a transformative role in enhancing patient outcomes and fostering greater trust in AI-powered healthcare solutions.

## Data Availability

This study did not generate any new data. All data discussed and analyzed are sourced from previously published research studies. The datasets referenced in the study have been fully described in Tables 8 and 9, including information on their variables, missing values, and the methods used in their original collection. The research remains a secondary analysis, utilizing existing datasets without the involvement of new data collection efforts

## CREDIT AUTHORSHIP CONTRIBUTION STATEMENT

Noor A. Aziz: Writing original draft (lead), Methodology (lead), Investigation (equal), Visualisation (equal), Formal analysis (lead), Writing review & editing (supporting), Data curation (lead). Awais Manzoor: Writing original draft (supporting), Investigation (equal), Visualisation (equal), Writing review & editing (supporting), Validation (supporting), Data curation (supporting). Muhammad Deedahwar Mazhar Qureshi: Writing review & editing (supporting), Writing original draft (supporting). M. Atif Qureshi: Conceptualisation (equal), Methodology (supporting), Writing original draft (supporting), Writing review & editing (lead), Validation (lead), Visualisation (equal), Supervision (lead), Project administration (lead), Investigation (supporting), Formal analysis (supporting). Wael Rashwan: Conceptualisation (equal), Methodology (supporting), Writing review & editing (supporting), Investigation (supporting), Validation (supporting), Supervision (supporting), Project administration (supporting).

## APPENDIX

**TABLE 1.** Summary of Database Searches.

| Name of Database | Search String |
| --- | --- |
| Scopus | TITLE-ABS-KEY ( ( ( explainable OR interpretable OR transparent OR accountable OR human-interpretable OR human-centered ) AND ( ai OR ml OR DL OR machine AND learning OR deep AND learning OR artificial AND intelligence OR computational AND intelligence ) OR XAI ) AND ( ( ( health* OR *medical OR clinical ) AND decision* AND ( support OR making OR aids ) AND system* ) OR cdss OR cds ) ) |
| Web of Science | AB=(( ( explainable OR interpretable OR transparent OR accountable OR human-interpretable OR human-centered ) AND ( ai OR ml OR DL OR machine AND learning OR deep AND learning OR artificial AND intelligence OR computational AND intelligence ) OR XAI ) AND ( ( ( health* OR medical OR clinical ) AND decision* AND ( support OR making OR aids ) AND system* ) OR cdss OR cds ) ) |
| Cochrane Library | ((Explainable OR Interpretable OR Transparent OR Accountable OR Human-interpretable OR Human-Centered) AND (AI OR ML OR DL OR machine learning OR Deep learning OR artificial intelligence OR computational intelligence) OR XAI) AND (((Health* OR *medical OR clinical) AND decision* AND (support OR making OR aids) AND system*) OR CDSS OR CDS) |
| PubMed | ((("Explainable"[Title/Abstract] OR "interpretable"[Title/Abstract] OR "Transparent"[Title/Abstract] OR "Accountable"[Title/Abstract] OR "Human-interpretable"[Title/Abstract] OR "Human-Centered"[Title/Abstract]) AND ("AI"[Title/Abstract] OR "ML"[Title/Abstract] OR "DL"[Title/Abstract] OR "machine learning"[Title/Abstract] OR "deep learning"[Title/Abstract] OR "artificial intelligence"[Title/Abstract] OR "computational intelligence"[Title/Abstract])) OR "XAI"[Title/Abstract]) AND (((("health*"[Title/Abstract] OR "medical"[Title/Abstract] OR "clinical"[Title/Abstract]) AND "decision*"[Title/Abstract] AND ("support"[Title/Abstract] OR "making"[Title/Abstract] OR "aids"[Title/Abstract]) AND "system*"[Title/Abstract]) OR "CDSS"[Title/Abstract] OR "CDS"[Title/Abstract]) |

**TABLE 2:**
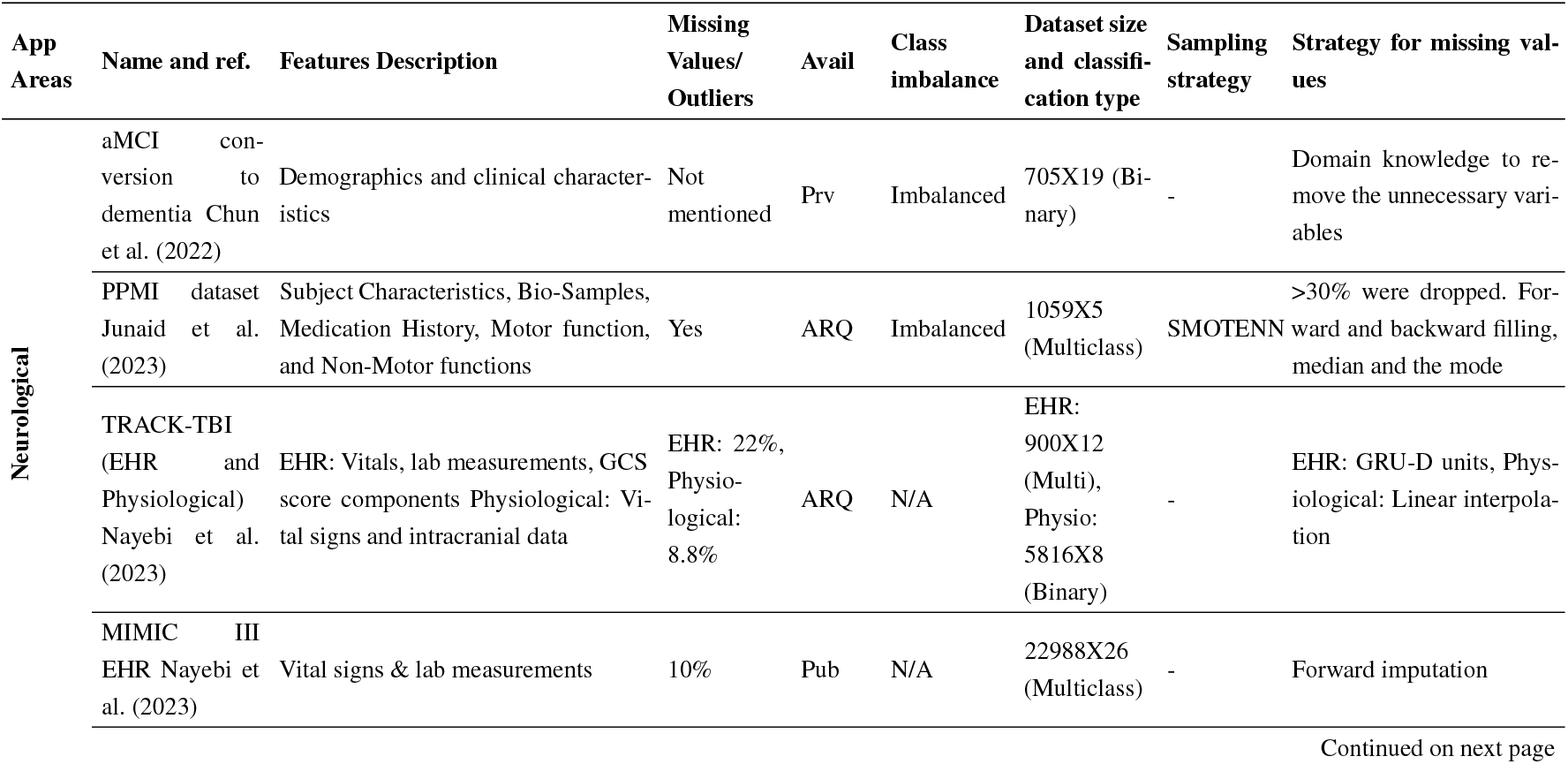
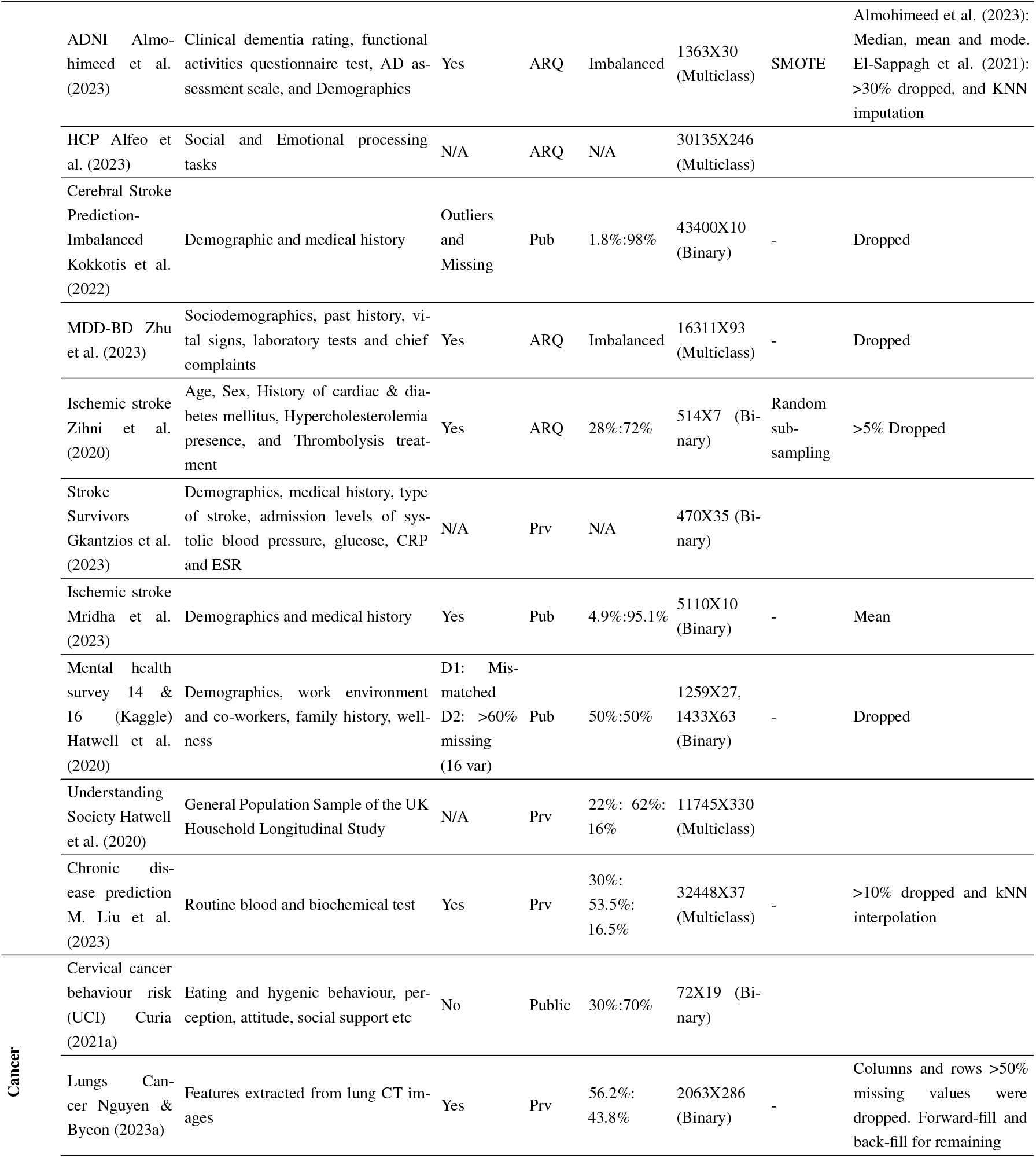
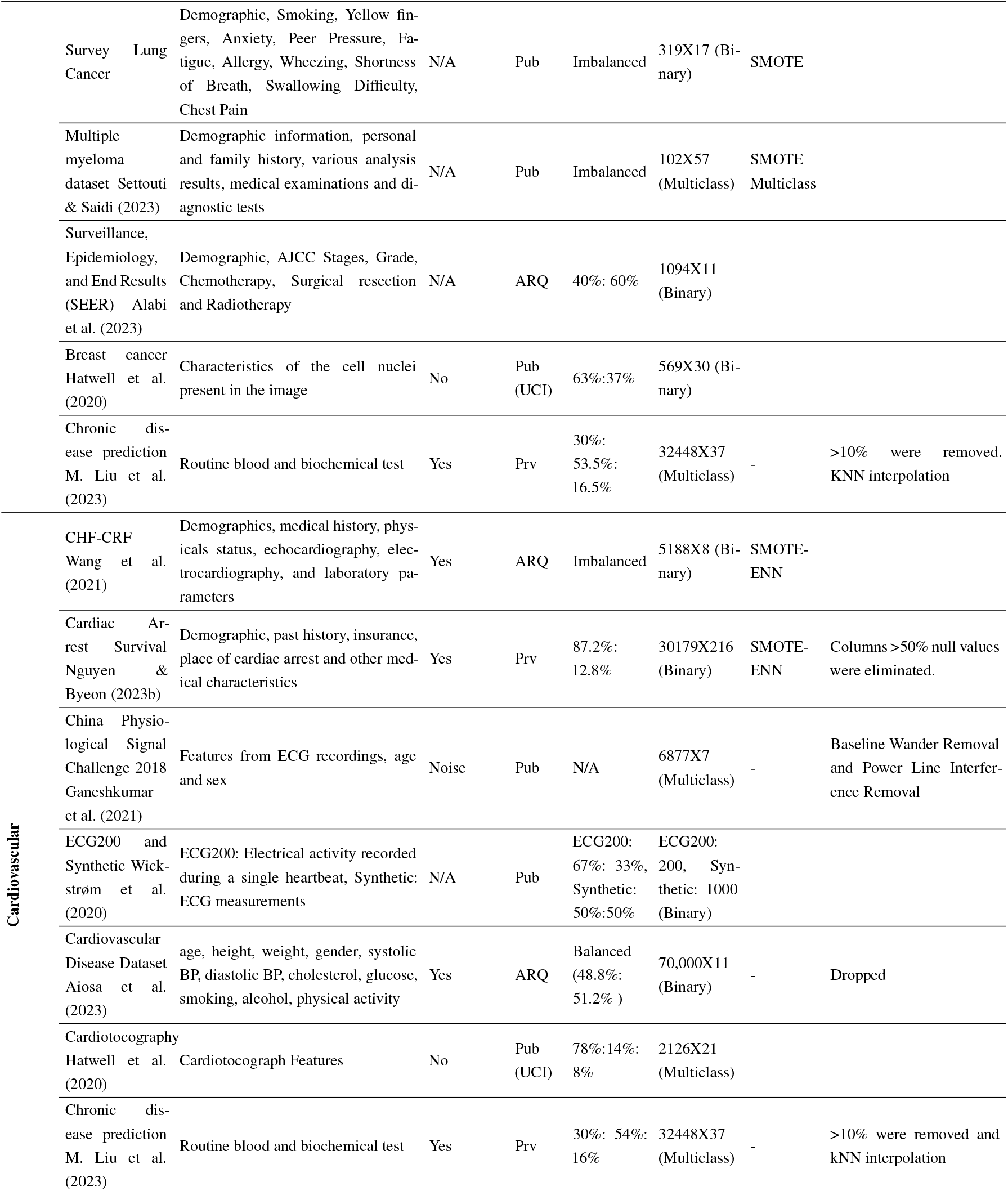
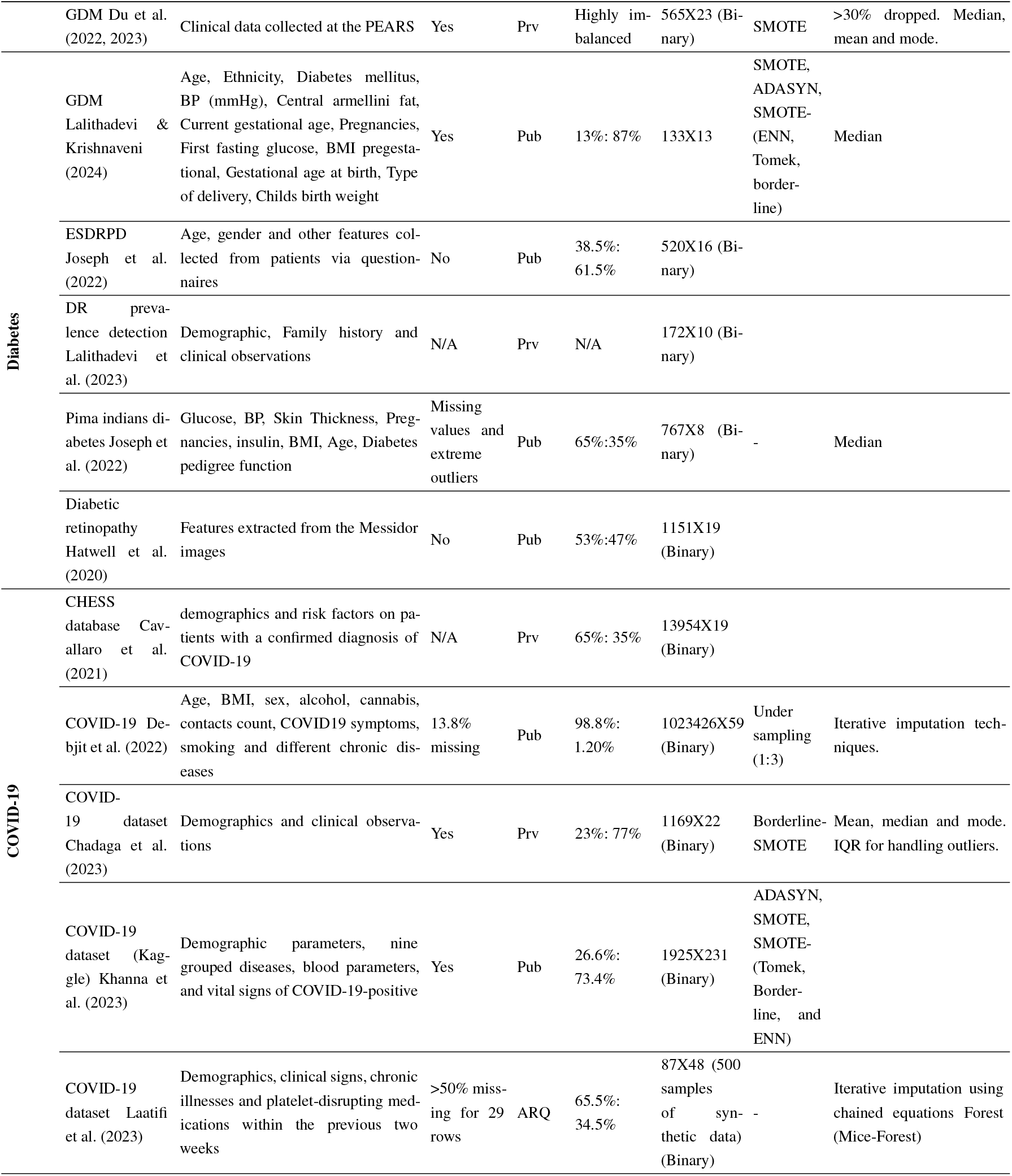
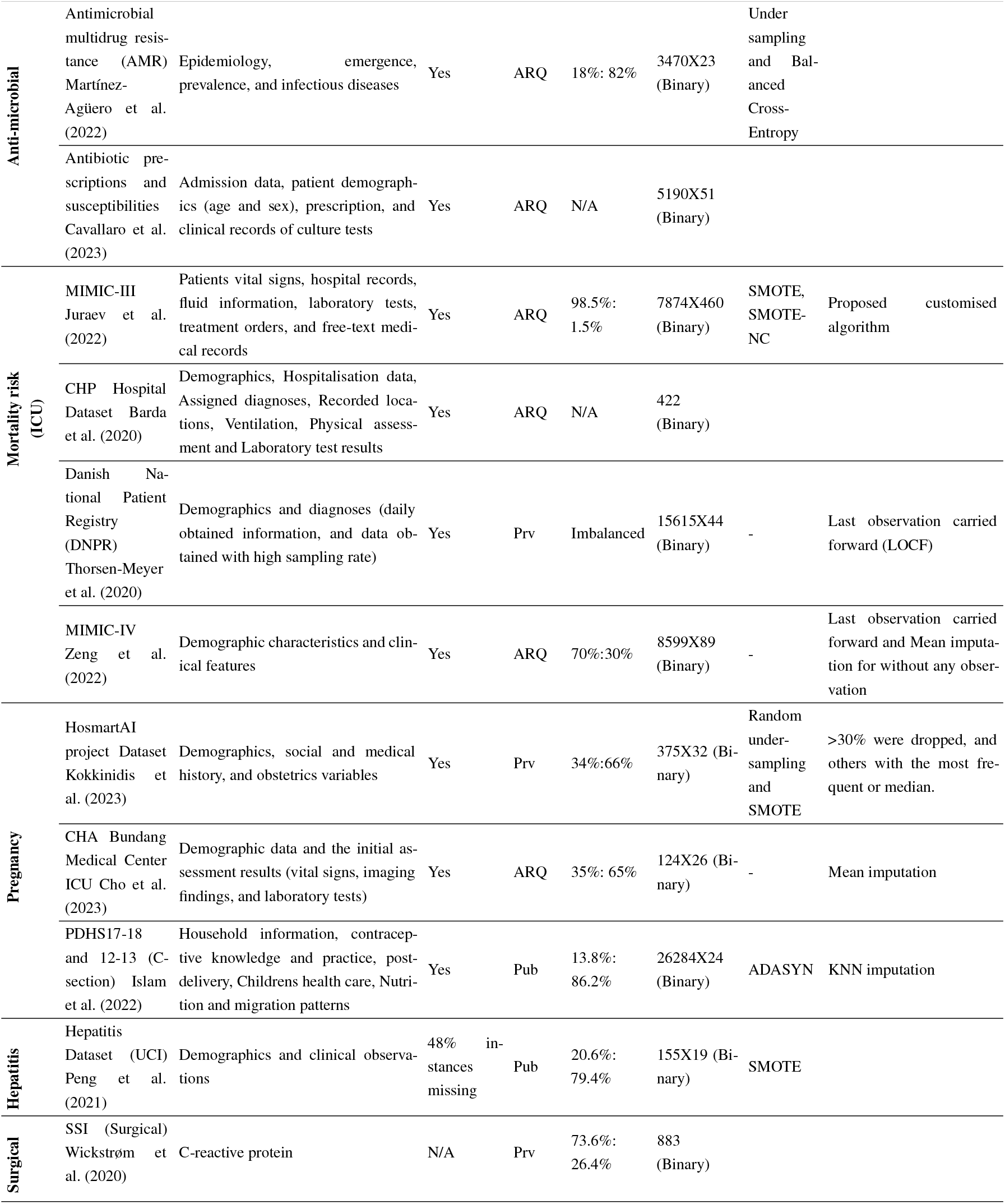
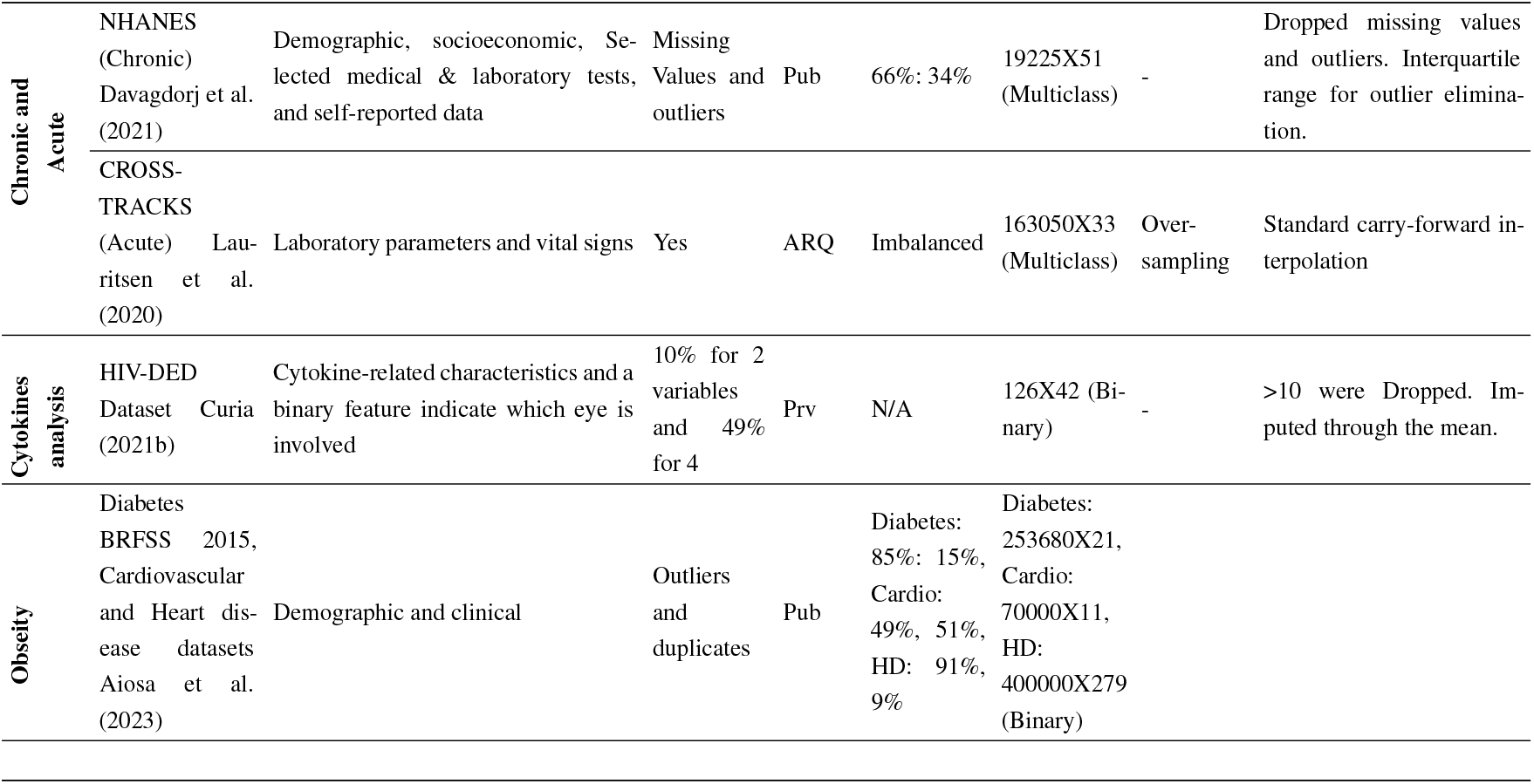
Overview of Tabular Datasets by Application Area, Availability, and Types of Data.

**TABLE 3:**
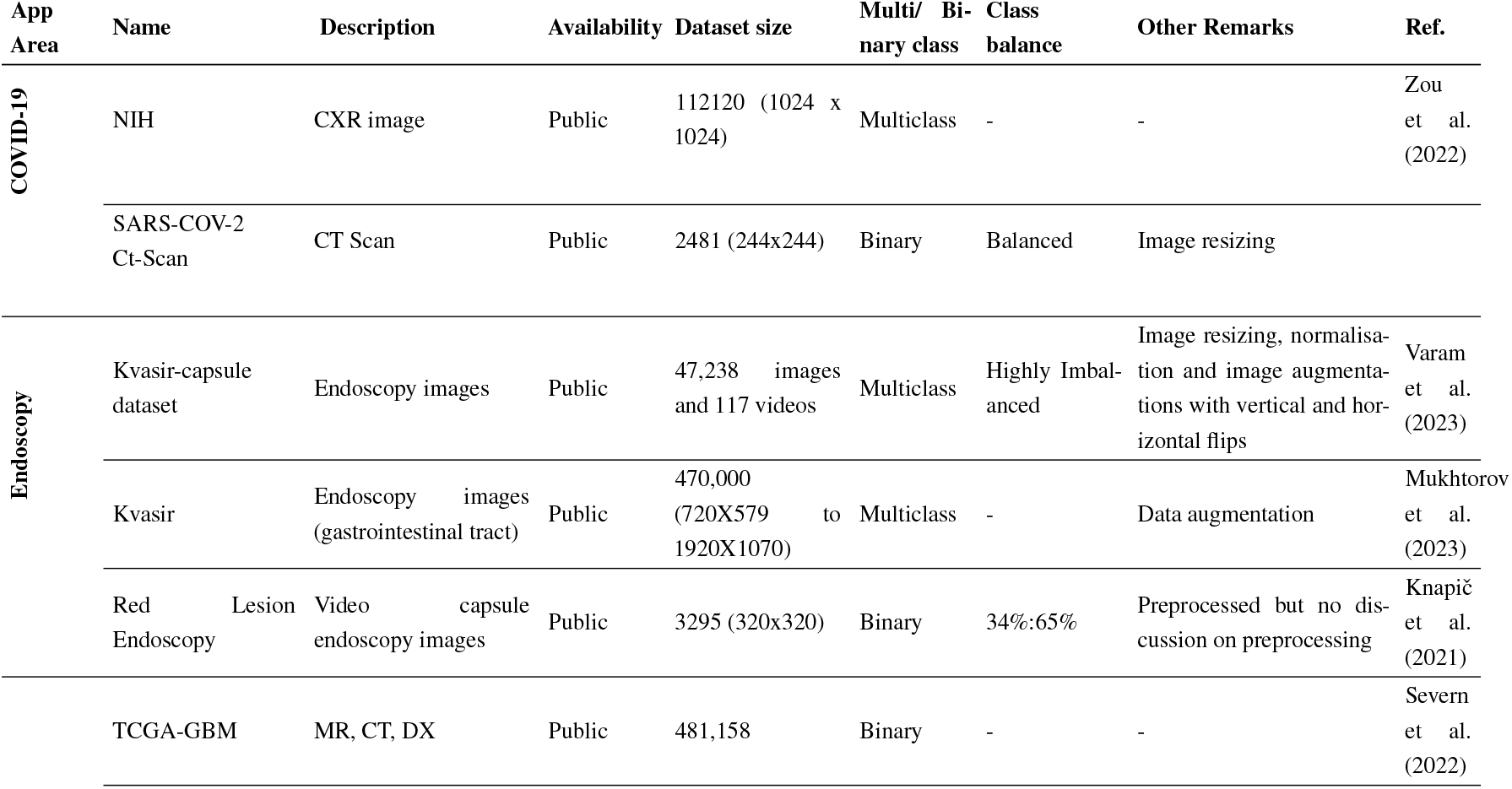
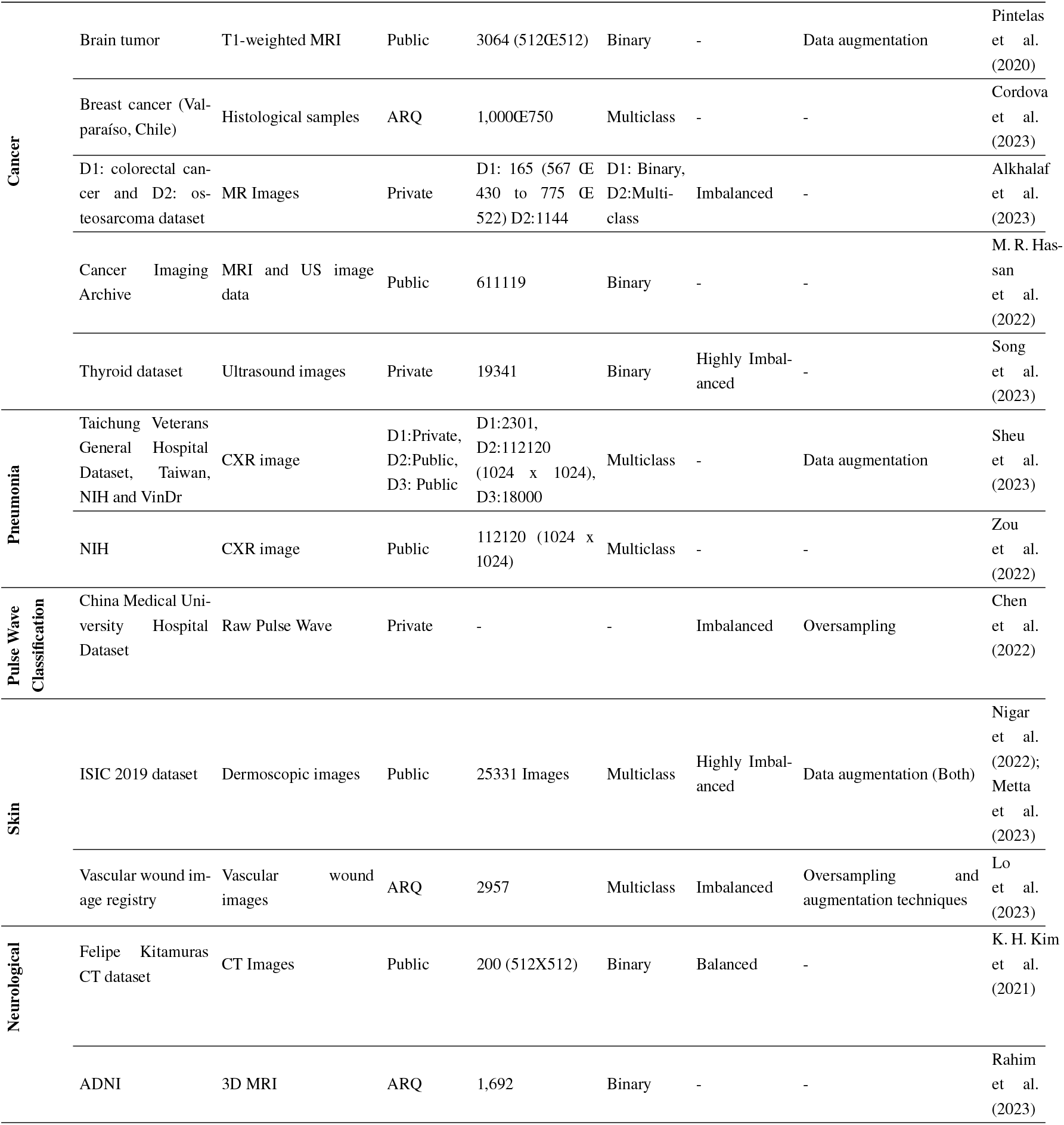
Overview of Image Datasets by Application Area, Availability, and Types of Data.

**TABLE 4:** Detailed Descriptions of Popular XAI Methods.

| XAI Method | Description | Applications in Healthcare |
| --- | --- | --- |
| **SHAP** | SHAP assigns each feature an importance value for a particular prediction by calculating Shapley values derived from cooperative game theory. This method ensures that the contribution of each feature is fairly distributed based on different feature combinations. | SHAP is widely used in healthcare to interpret predictions from complex models, particularly in cases like patient risk stratification, diagnosis, and treatment recommendations. It is applicable to both tabular and imaging data. |
| **LIME** | LIME approximates the model locally by generating a new dataset consisting of permuted samples and their corresponding predictions. It then trains an interpretable model on this new dataset to explain the original models predictions for individual instances. | LIME is popular in scenarios requiring localized explanations, such as understanding why a specific patient's outcome was predicted in clinical settings. This makes it useful for personalized medicine and decision-making. |
| **GradCAM** | GradCAM produces a coarse localisation map by computing the gradient of the target class score concerning the feature maps of the last convolutional layer. This map highlights the important regions in the input image that are most influential in the models decision. | GradCAM is predominantly used in medical imaging to interpret CNNs' decisions, aiding in diagnosing diseases such as cancer by highlighting critical regions in medical images like MRI or CT scans. |
| **Anchors** | Anchors are high-precision rules that explain the decision of any classifier. An anchor is a set of conditions that sufficiently anchors a prediction, meaning that changes to other features will not affect the outcome as long as the anchor conditions hold. | Anchors are effective in rule-based explanations. They are often used in clinical decision support systems to explain why a specific decision or recommendation was made, making them useful in complex clinical scenarios. |
| **Saliency Maps** | Saliency maps highlight the pixels in an input image that impact the models predictions most, typically by calculating the output gradient concerning the input image. | Saliency maps are used in medical imaging to identify areas of an image that are most relevant to a model's decision, such as locating tumours or other anomalies in radiological scans. |
| **ELI5** | ELI5 is a library that provides a unified interface for various explanation methods, such as feature importance, and offers a simple way to debug ML models by understanding which features influence the outcome. | ELI5 is used for debugging and understanding ML models in healthcare, helping clinicians and researchers understand which features are driving predictions in models used for patient diagnosis or treatment plans. |
| **GradCAM++** | An extension of GradCAM, GradCAM++ uses a weighted combination of the positive partial derivatives of the last convolutional layers feature maps, which allows it to produce more accurate and detailed class activation maps compared to GradCAM. | GradCAM++ is particularly useful in situations where multiple objects are present in the image and helps to refine the interpretation of complex medical images by providing more detailed localisation. |
| **TabNet** | TabNet is a deep learning model tailored for tabular data. It uses sequential attention to select features at each decision step, making it both interpretable and highly accurate. | TabNet is utilised in healthcare for analysing tabular data, like predicting patient outcomes from EHR, delivering both predictions and explanations in one model. |
| **PDP** | PDPs show the relationship between a feature and the predicted outcome while keeping all other features constant. This helps in understanding how changes in a feature affect the prediction. | PDPs are useful in healthcare for analysing how factors like blood pressure and cholesterol levels affect the risk of heart disease. |
| **WindowSHAP** | A variant of SHAP, WindowSHAP is tailored for time-series data. It calculates Shapley values for different time windows, helping to identify which time intervals are most important for a models prediction. | WindowSHAP is used in healthcare to interpret models predicting outcomes from time-series data, such as patient monitoring systems that track vital signs over time. |
| **ICE** | ICE plots visualize the relationship between a feature and the predicted outcome for individual instances rather than averaging across all instances like PDPs. This provides more detailed insights into the models behaviour for specific cases. | ICE plots are applied in personalised medicine, where they help understand how changing a particular feature (e.g., medication dosage) might affect individual patients' outcomes. |
| **LRP** | LRP decomposes the prediction of a neural network by propagating the prediction backwards through the network, redistributing the prediction score to the input features in a way that highlights the most relevant features. | LRP is used in deep learning models for tasks like medical image classification, helping to explain which parts of the input image contributed most to the models decision. |

## Footnotes

‡ 451, 351, 229, and 195 results from Scopus, Web of Science, Cochrane Library, and PubMed, respectively.

§ added during the forward and backward chaining process.

¶ supervisor author of the students

\# supervisor authors

